# Effects of Reimbursement Policy Change on Utilization of Injectable Semaglutide for Type 2 Diabetes and on Glycemic Control Among Discontinuers: A Danish Nationwide Cohort Study

**DOI:** 10.64898/2026.09.23.26363640

**Authors:** Emma Simonsen, Martin Thomsen Ernst, Diana Hedevang Christensen, Helene Kildegaard, Jens Søndergaard, Reimar W. Thomsen, Anton Pottegård

## Abstract

**Objectives:** To evaluate the impact of reimbursement restrictions for glucagon-like peptide-1 receptor agonists (GLP-1RAs) for type 2 diabetes, introduced in Denmark in November 2024, on injectable semaglutide (Ozempic®) utilization patterns and subsequent HbA1c changes among discontinuers.

**Design:** Nationwide registry-based cohort study.

**Setting:** Denmark.

**Participants:** Adults treated with injectable semaglutide for diabetes (January 2018 to December 2025).

**Main outcome measures:** We described temporal patterns in injectable semaglutide use before and after the policy change. Among semaglutide continuers after the policy change, we assessed whether continuation met a key criterion in the new policy: non-feasibility of sodium-glucose cotransporter 2 inhibitor use and high cardiovascular risk. We characterized semaglutide discontinuers around the policy implementation (March 2024 to November 2024) and subsequent treatment switches. Among discontinuers, we evaluated HbA1c changes and performed interrupted time series (ITS) analyses.

**Results:** Following the policy change, use of injectable semaglutide declined modestly before subsequently increasing. Among 61,120 individuals classified as semaglutide continuers, 27,143 (44%) were already using an SGLT2i. Of the remaining 33,977 (56%), only 1,862 (5.5%) met a key reimbursement criterion (i.e., non-feasibility of SGLT2i use and high cardiovascular risk). Among 79,305 individuals with a semaglutide fill between March 2024 and November 2024, 22,096 (28%) were classified as discontinuers. Within six months, ∼40% of discontinuers switched to other glucose-lowering therapies, most commonly SGLT2is (24%). 37% of discontinuers later reinitiated semaglutide. Among individuals with an on-treatment HbA1c measurement (within 12 months before their last semaglutide fill) and an off-treatment HbA1c measurement (6-12 months after their last semaglutide fill) (n=13,017), mean HbA1c increased from 49 to 57 mmol/mol (mean change +7.4 mmol/mol, 95% CI 7.2-7.6). ITS analysis showed that HbA1c increased by 5.9 mmol/mol (95% CI 5.1-6.7) at six months post-discontinuation, which attenuated to 3.1 mmol/mol (95% CI 2.2-4.1) at 12 months; raw monthly means (not included in the ITS model) suggested this persisted through 18 months.

**Conclusions:** Following the policy change, population-level use of injectable semaglutide declined modestly before increasing again. Most discontinuers did not initiate another glucose-lowering therapy and experienced an increase in HbA1c that did not fully return to pre-discontinuation levels during follow-up.

**What is already known on this topic:**

- The rapid uptake and high costs of GLP-1 receptor agonists, including injectable semaglutide for diabetes (Ozempic®), have prompted health systems to consider how access to these treatments should be prioritized and reimbursed.
- Little is known about how reimbursement restrictions on GLP-1 receptor agonists influence treatment patterns and glycemic control after treatment discontinuation.

**What this study adds:**

- Following a national reimbursement restriction in Denmark, more than one in four patients discontinued injectable semaglutide for diabetes, and among those who switched treatment, SGLT2 inhibitors were the most common alternative, while most discontinuers did not initiate another glucose-lowering drug within six months. Approximately one in three discontinuers reinitiated their semaglutide therapy more than six months after stopping.
- Discontinuation of injectable semaglutide for diabetes was followed by a rise in HbA1c that, despite improvement over time, did not return to pre-discontinuation levels over 18 months, highlighting a potential clinical consequence of restricting access to an effective glucose-lowering treatment option.

## Introduction

Glucagon-like peptide-1 receptor agonists (GLP-1RAs), including injectable semaglutide (Ozempic®), have become a cornerstone of treatment for type 2 diabetes (T2D) but have also contributed to rising healthcare expenditures. In response to increasing utilization and escalating costs, Denmark’s national health authorities introduced more restrictive national reimbursement criteria for GLP-1RAs in November 2024.^1^ The nationwide implementation of these reimbursement criteria created a natural experiment for evaluating reimbursement policies in healthcare systems facing similar challenges in balancing access to high-cost therapies with budgetary constraints.

Previously, reimbursement for GLP-1RAs in Denmark was broadly available to patients with T2D who had not achieved adequate glycemic control despite treatment with metformin plus at least one additional non-GLP-1RA glucose-lowering drug, or for whom additional treatment with glucose-lowering drugs other than GLP-1RAs was considered inappropriate.^2^ Under the revised national policy, the Danish Medicines Agency restricted reimbursement to two groups with T2D. First, patients who could not use sodium-glucose cotransporter 2 inhibitors (SGLT2is) due to intolerance or severe renal impairment, and who either had (1) cardiovascular or renal disease (regardless of glycemic control) or had (2) multiple cardiovascular risk factors with inadequate glycemic control on metformin. Second, patients who had not achieved adequate glycemic control despite treatment with *all relevant* oral glucose-lowering drugs, including SGLT2is.^3^ Consequently, the policy may have led many patients with T2D to discontinue their GLP-1RA treatment and switch to alternative drugs, with potential implications for diabetes management. GLP-1RAs and SGLT2is are both effective glucose-lowering drugs that reduce cardiovascular events and mortality; however, their profiles differ in clinically meaningful ways.^4–7^ SGLT2is seem superior in preventing heart-failure hospitalizations and slowing renal-disease progression, whereas GLP-1RAs appear to be more effective in reducing stroke risk,^4–6^ but large head-to-head trials are lacking.^4–7^ Although GLP-1RAs provide greater HbA1c reductions and slightly more weight loss than SGLT2is at T2D-indicated doses,^8,9^ SGLT2is remain recommended alternatives to GLP-1RAs.^10^

Since injectable semaglutide accounted for roughly 85% of GLP-1RA use for T2D in Denmark during the study period,^11^ we focused on patients treated with injectable semaglutide. Specifically, we addressed two complementary questions. First, for patients who continued treatment with semaglutide, we assessed whether continuation was consistent with the reimbursement policy by operationalizing the first reimbursement criterion, i.e., non-feasibility of SGLT2i use and high cardiovascular risk. Second, we assessed the number of semaglutide-treated patients who discontinued their treatment and compared them with those who did not, with respect to baseline patient characteristics and prior treatment patterns. Among patients who changed treatment, we described the therapies they switched to and evaluated the impact on glycemic control by analyzing within-person HbA1c changes, with and without semaglutide treatment. Together, these analyses provided insight into the implementation of a new national reimbursement policy and its short-term consequences.

## Methods

### Setting

Denmark has a tax-funded healthcare system in which most prescription drugs are partially reimbursed through a national tiered reimbursement scheme. Because reimbursement criteria are set nationally, changes to them may affect prescribing patterns across the country. Following an extended public consultation process that began in 2022,^12^ the Danish Medicines Agency, guided by the Danish Medicines Council, announced its official revision of the reimbursement criteria for GLP-1RAs, initiating a transition period before the revised criteria were formally implemented and became fully effective on 25 November 2024.^13^ Under the new reimbursement policy, eligibility for reimbursed GLP-1RA treatment became substantially more restrictive.

### Data sources

We used data from the Danish National Prescription Registry, which includes information on dispensing dates, prescribers, and Anatomical Therapeutic Chemical (ATC) drug classifications.^14^ To capture patients’ broader glucose-lowering drug use and characterize switches in therapy, we additionally retrieved all fills of glucose-lowering drugs recorded in the registry from 1995 onward. The Civil Registration System was used to collect demographic information, including date of birth and sex.^15^ The Danish National Patient Registry was used to collect data on in- and outpatient hospital contacts coded according to the International Classification of Diseases, Tenth Revision (ICD-10).^16^ The Danish Register of Laboratory Results was used to collect laboratory data, including HbA1c values.^17^ All data sources were linked using the unique personal civil registration number assigned to all Danish residents.^15^

### Study population and exposure

On 8 February 2018, injectable semaglutide for diabetes management (Ozempic®) received EU-wide marketing authorization (including Denmark as an EU member state). We identified all adults aged at least 18 years or older who filled a first prescription for semaglutide from 1 January 2018 (calendar year of market entry in Denmark) to 31 December 2025 (end of prescription data availability). To include only patients with T2D, individuals were required to have at least one fill for an additional glucose-lowering drug class within one year before their first semaglutide fill. Only individuals with at least one year of continuous look-back before their first semaglutide fill (index date) were included to allow capture of prior medical history and medication use. Individuals were followed from their first semaglutide fill until death, emigration, or 31 December 2025. From this source population, we first described temporal patterns in glucose-lowering treatment use (2018–2025) and glycemic monitoring one year before and after the policy change became fully effective on 25 November 2024. Hereafter, we constructed analytic sub-cohorts for assessment of reimbursement-consistent semaglutide continuation and, among discontinuers, switching and changes in HbA1c (**Figure 1**).

**Figure 1.**
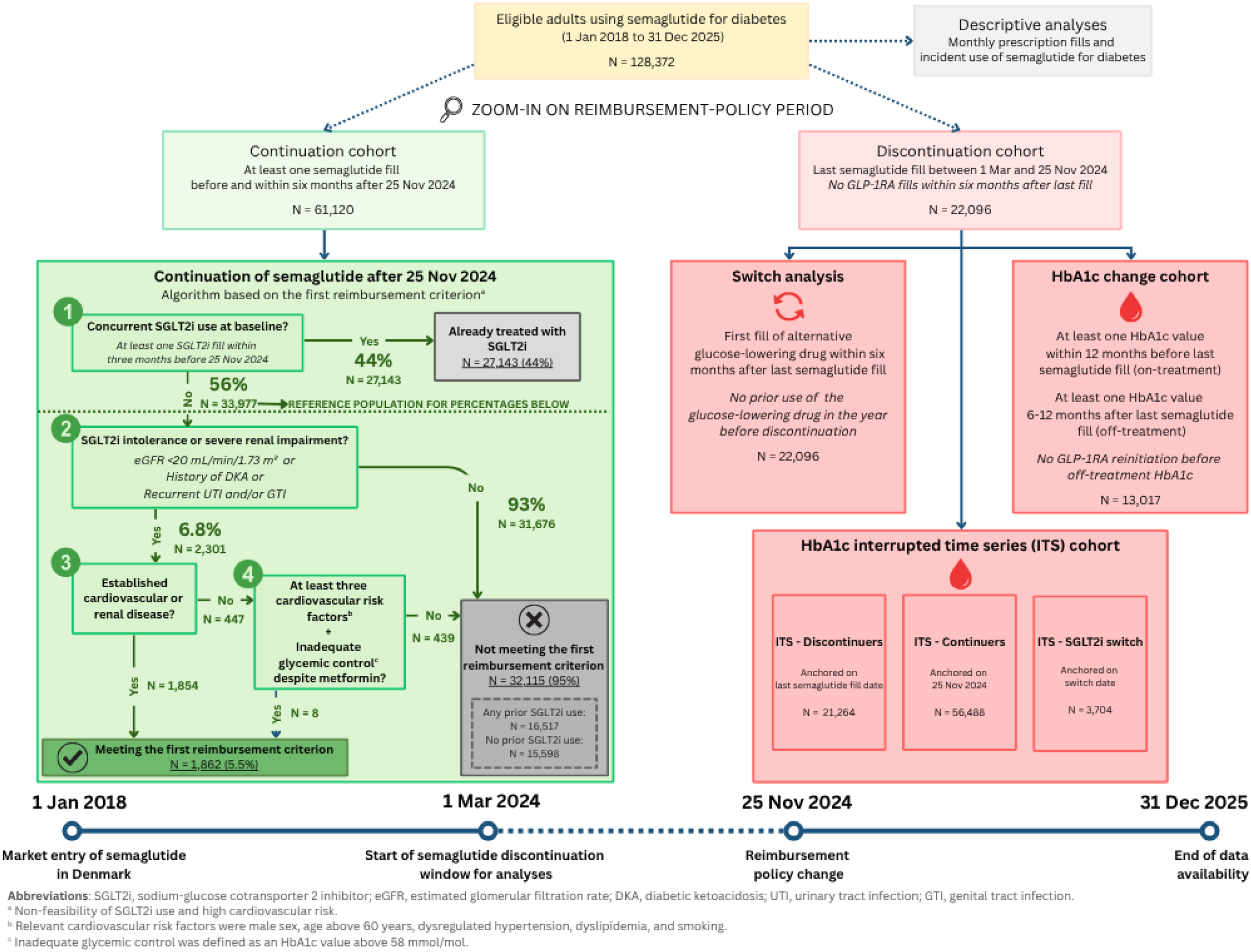
Flow diagram of the study population and analytic cohorts. Eligible adults using semaglutide for diabetes management from January 2018 through December 2025 were identified. Individuals were subsequently classified into analytic cohorts for continuation, discontinuation (including switching), and HbA1c analyses.

### Glucose-lowering drug use (2018–2025) and HbA1c monitoring one year before/after policy change

We first described glucose-lowering treatment patterns from injectable semaglutide market entry and glycemic monitoring around the reimbursement change. These analyses provided a temporal context for interpreting treatment switches and HbA1c changes. Specifically, we calculated (i) the monthly number of fills of semaglutide and other glucose-lowering drugs from 1 January 2018 to 31 December 2025, ii) the monthly rate of initiators of semaglutide and other glucose-lowering drugs, i.e., first filled prescriptions of these drugs following a five-year washout period, from 1 January 2018 to 31 December 2025, and (iii) the number of HbA1c measurements per quarter during the one year before and after the policy change, to characterize treatment use and glycemic monitoring patterns surrounding the policy implementation.

### Characterization of reimbursement-consistent semaglutide continuation

We developed a prespecified algorithm to classify whether continued injectable semaglutide use after the policy change became fully effective on 25 November 2024 was consistent with the first reimbursement criterion operationalized using registry data (**Figure 1**). Our algorithm was operationalized to capture the first reimbursement criterion only, i.e., patients with SGLT2i intolerance or severe renal impairment, who either had (1) cardiovascular/renal disease or (2) multiple cardiovascular risk factors with inadequate glycemic control on metformin. The second reimbursement criterion could not be operationalized in our registries, as it required individualized clinical assessment of whether “adequate” glycemic control had been achieved despite prior use of “all relevant” oral glucose-lowering drugs. This could not be reliably captured using registry data alone, as individualized HbA1c targets and their determinants, such as frailty, cohabitation status, occupational risks related to hypoglycemia, etc., were not available.

To operationalize the first criterion, we applied a stepwise algorithm. First, we identified individuals with concurrent SGLT2i use at baseline, defined as at least one SGLT2i fill within three months before 25 November 2024. These individuals were excluded from consideration as potential candidates for switching to an SGLT2i. Second, among the remaining individuals, we identified those with likely SGLT2i intolerance or severe renal impairment, including the most recent estimated glomerular filtration rate <20 mL/min/1.73 m² within two years before the policy change (sensitivity thresholds: <25 and <30 mL/min/1.73 m²), any history of diabetic ketoacidosis, and recurrent urinary or genital infections, defined as at least two urinary and/or genital infection events in the 12 months before the policy change. For urinary and genital infections, events were identified from both hospital diagnoses and community pharmacy prescription fills, e.g., antibiotic and antifungal medications. To increase the likelihood that diagnoses and prescription fills represented distinct infection events, consecutive events were required to be separated by at least one month. Third, within the subgroup of individuals with SGLT2i intolerance or severe renal impairment, individuals were classified in a two-step process. We first identified those with established cardiovascular or renal disease. Cardiovascular disease was defined as any hospital diagnosis of ischemic heart disease, cerebrovascular disease, atherosclerotic peripheral vascular disease, or heart failure (**Table S2**). Renal disease was defined as any hospital diagnosis indicating impaired renal function (including acute renal failure and chronic renal disease), any dialysis procedure, and persistent kidney dysfunction defined as either eGFR <60 mL/min/1.73 m² based on the most recent measurement (excluding inpatient values) or urine albumin-creatinine ratio ≥30 mg/g measured on at least two occasions at least three months apart (**Table S2**). Among the remaining individuals, we identified those with both (i) at least three cardiovascular risk factors (male sex, age >60 years, dysregulated hypertension [>130/80 despite treatment], dyslipidemia [LDL-cholesterol >1.8 mmol/l despite statin treatment, defined as a statin fill within the three months preceding the measurement], and smoking history of more than 10 pack-years)^18^ and (ii) inadequate glycemic control despite metformin, defined as an HbA1c >58 mmol/mol based on the most recent measurement within the six months before 25 November 2024 (sensitivity thresholds: >48, >53, >64, and >70 mmol/mol) and at least one metformin fill within the three months before 25 November 2024. Due to the absence of blood pressure and smoking status in the registries, dysregulated hypertension was operationalized as the use of at least two concurrent antihypertensive drug classes, and smoking exposure was operationalized as the presence of registry-based smoking indicators (**Table S2**). Individuals not meeting these conditions were assigned to the residual group. In a post hoc analysis, we repeated the algorithm assuming that any prior use of an SGLT2i before the policy change represented SGLT2i intolerance, thereby allowing these individuals to meet the first reimbursement criterion if all other algorithm components were fulfilled.

### Potential reimbursement-related semaglutide discontinuation

Based on an exploratory descriptive analysis of injectable semaglutide discontinuation patterns conducted as part of the study protocol, the period from March 2024 to November 2024 was selected to define potential reimbursement-related semaglutide discontinuation (**Figure S1**). Accordingly, semaglutide discontinuation was defined as the last fill during this period with no subsequent GLP-1RA fills within the following six months. We compared baseline patient characteristics and prior treatment patterns between individuals discontinuing semaglutide during this period and those who continued treatment. Among individuals discontinuing semaglutide, we examined subsequent treatment switching and changes in HbA1c (**Figure 1**).

### Analysis of treatment switches

To characterize treatment changes following semaglutide discontinuation, we described switches from injectable semaglutide to alternative glucose-lowering drugs. A switch was defined as the first fill of SGLT2i, dipeptidyl peptidase-4 inhibitor (DPP4i), sulfonylurea (SU), metformin, insulin, or glitazone occurring within six months of semaglutide discontinuation, provided that the individual had no fills of that glucose-lowering drug in the year before discontinuation. We estimated the proportions of semaglutide users who switched to each alternative glucose-lowering drug class, both overall and stratified by the baseline glucose-lowering regimen used during the three months before the switch. We also assessed reinitiation of semaglutide among discontinuers, defined as a semaglutide fill occurring more than six months after discontinuation. We also conducted subgroup analyses stratified by duration of semaglutide use, last dose of semaglutide, and baseline HbA1c. HbA1c was categorized using thresholds based on guideline-relevant ranges,^19^ using the most recent HbA1c measurement within the six months before the last Ozempic fill.

### Analysis of HbA1c changes

For the HbA1c analyses, we required an ‘on-treatment’ HbA1c value measured after initiation of injectable semaglutide and within the 12 months before the last semaglutide fill, an ‘off-semaglutide treatment’ HbA1c value within 6–12 months after the last injectable semaglutide fill, and no GLP-1RA reinitiation before the off-treatment HbA1c measurement. These analyses included all adults who discontinued injectable semaglutide, irrespective of whether they subsequently switched to another glucose-lowering therapy, provided they met these HbA1c measurement requirements. We ignored the first three months because HbA1c reflects average glycemic control over the preceding three months and would therefore not yet indicate effects of semaglutide cessation, and the subsequent 6–12-month window allowed sufficient time for treatment effects to dissipate, including any residual effects due to delayed medication depletion or stockpiling.

The primary outcome was the within-individual change in HbA1c, defined as the difference between the most recent on-treatment HbA1c and the first eligible off-treatment HbA1c, using a paired t-test. A similar paired comparison was made using the most recent off-semaglutide treatment HbA1c value available to assess longer-term changes. To aid interpretation of the within-individual changes, the time elapsed between the last Ozempic fill and the most recent HbA1c measurement was described (median and interquartile range).

To aid interpretation, we described the time from the last injectable semaglutide fill to the most recent off-semaglutide treatment HbA1c value, and performed predefined subgroup and sensitivity analyses, including stratification by patient characteristics, exclusion of potential stockpiling, and complementary analyses of patients switching to SGLT2is (supplemental methods).

### Complementary analyses of HbA1c changes

As a complementary analysis, we conducted interrupted time series analyses^20^ to assess changes in HbA1c over time following semaglutide discontinuation. A segmented linear regression model was used to estimate changes in level and slope (i.e., trend over time) following the interruption date. Unlike the within-person, paired HbA1c analyses, the interrupted time series analyses were not restricted to individuals meeting the paired measurement requirements. Following a post hoc extension of laboratory data availability, HbA1c measurements were available through 5 February 2026 and were used to describe levels beyond the 12-month analysis window; these measurements were not included in the interrupted time series analyses.

We applied the same interrupted time series approach to adults who continued filling injectable semaglutide after the policy change, using 25 November 2024 as the interruption date. For discontinuers, sensitivity analyses excluded the three months immediately following the discontinuation date, and a parallel interrupted time series model was conducted among SGLT2i switchers, anchored on the switch date. For individuals who discontinued semaglutide, the interruption date was defined as the individual’s date of discontinuation.

All analyses were performed using R version 4.4.3.

### Study transparency and reproducibility^21^

The study protocol was registered at the Real-World Evidence Registry (https://osf.io/jbzda) before the commencement of any analyses, with subsequent amendments also made available (https://osf.io/gxhp3). Detailed methodological information, including all codes and definitions used, is available in the protocol and its amendments. Due to Danish data protection regulations, individual-level data cannot be shared directly by the authors. De-identified data can be made available for authorized researchers after an application to ForskerService at the Danish Health Data Authority. The source code used for data management and statistical analyses is available at (https://gitlab.sdu.dk/pharmacoepi/Ozempic-Reimbursement). The manuscript is aligned with the REporting of studies Conducted using Observational Routinely-collected health Data (RECORD) statement.^22^

## Results

A total of 152,965 individuals filled at least one prescription for injectable semaglutide for diabetes management during the study period from 1 January 2018 to 31 December 2025. After restricting to adults, requiring at least one year of available look-back, and prior use of glucose-lowering drugs, 128,372 individuals remained and constituted the main study population of semaglutide users with T2D.

From the main study population (N = 128,372), several partially overlapping analytic sub-cohorts were identified, including 61,120 in the semaglutide continuation cohort who filled a prescription before and six months after 25 November 2024, 22,096 individuals in the discontinuation cohort, 13,017 in the HbA1c change cohort, and three HbA1c interrupted time series cohorts comprising 21,264 discontinuers, 56,488 continuers, and 3,704 SGLT2i switchers (**Figure 1**).

Among the 79,305 individuals with a semaglutide fill between March and November 2024, 57,209 individuals continued treatment through 25 November 2024, whereas 22,096 individuals discontinued treatment. Compared with individuals who continued semaglutide treatment, those who discontinued were more often female and had better glycemic control at baseline (HbA1c 46 vs. 51 mmol/mol). Discontinuers were also less likely to have established cardiovascular disease, including ischemic heart disease and heart failure, or to have received an SGLT2i before, with detailed baseline characteristics shown in **Table S3**.

### Glucose-lowering drug use (2018–2025) and HbA1c monitoring one year before/after policy change

Across the full study period, the number of monthly injectable semaglutide fills increased steadily following market entry in 2018. In the months up to the policy change in November 2024, semaglutide fills showed a modest decline, before increasing gradually in the months following the policy change (**Figure 2**). In contrast, fills for other glucose-lowering drug classes remained relatively stable over time. Overall, semaglutide accounted for a substantial proportion of overall glucose-lowering drug fills, particularly from 2023 onward. **Figure 3** shows the monthly incidence rate of filled prescriptions for glucose-lowering drug classes, where incidence was defined as the first filled prescription in five years. Incident injectable semaglutide prescribing peaked in early 2023, declined sharply during the second half of 2023, and increased briefly before the November 2024 policy change. It then fell again around the time of the policy change and rose once more in mid-2025. Incident prescribing of both SGLT2is and DPP4is increased around the November 2024 policy change before declining again in 2025.

**Figure 2.**
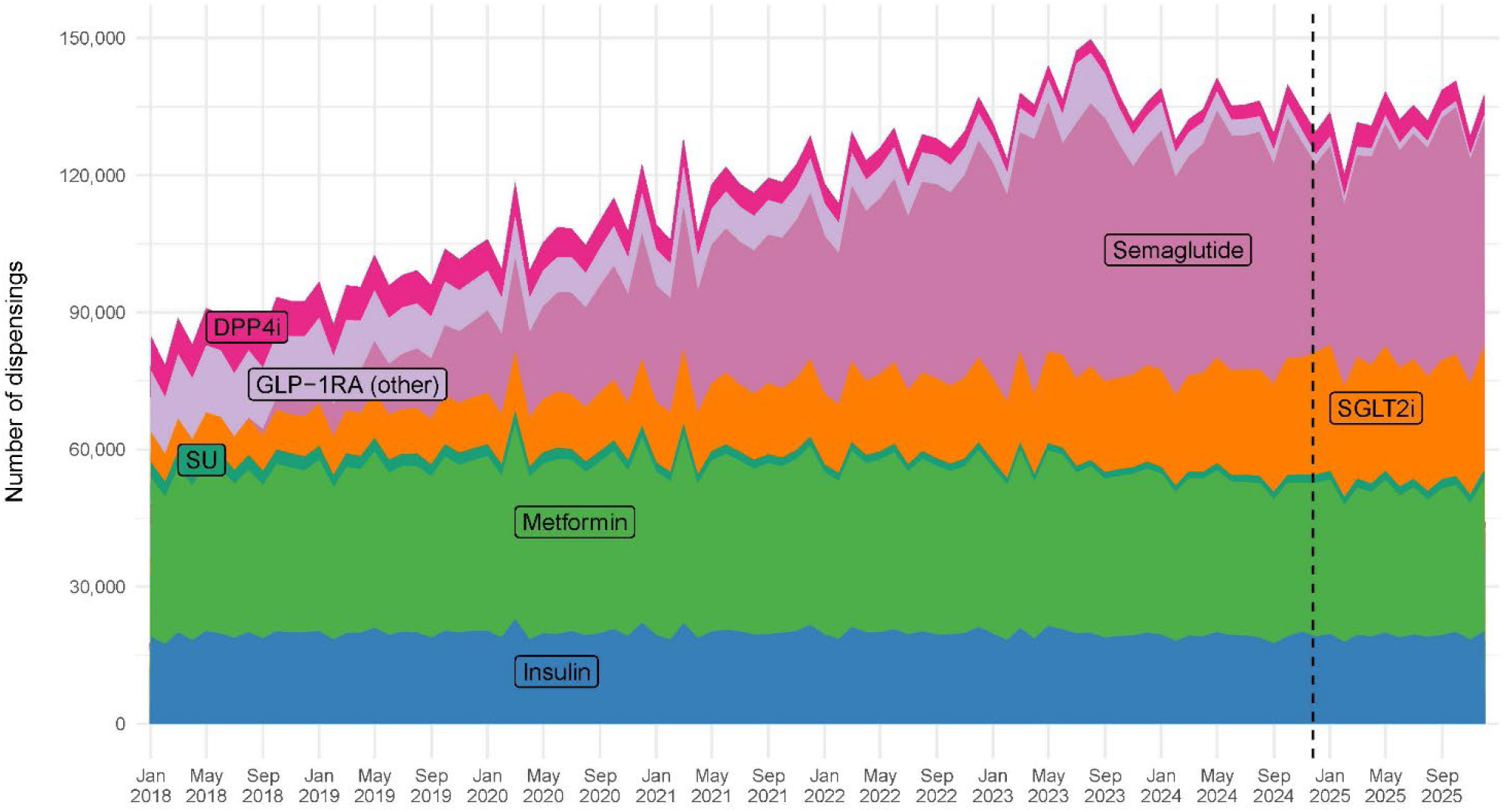
Monthly number of filled prescriptions for glucose-lowering drug classes in Denmark from January 2018 to December 2025 among eligible adults with at least one semaglutide prescription redemption in the period. Trends are shown for semaglutide, ‘other’ glucagon-like peptide-1 receptor agonists (GLP-1RA other; including Rybelsus® and Victoza®), sodium-glucose cotransporter 2 inhibitors (SGLT2i), dipeptidyl peptidase-4 inhibitors (DPP4i), sulfonylureas (SU), metformin, and insulin; glitazones are not visible due to very low prescription counts. The dashed line indicates the policy change on 25 November 2024.

**Figure 3.**
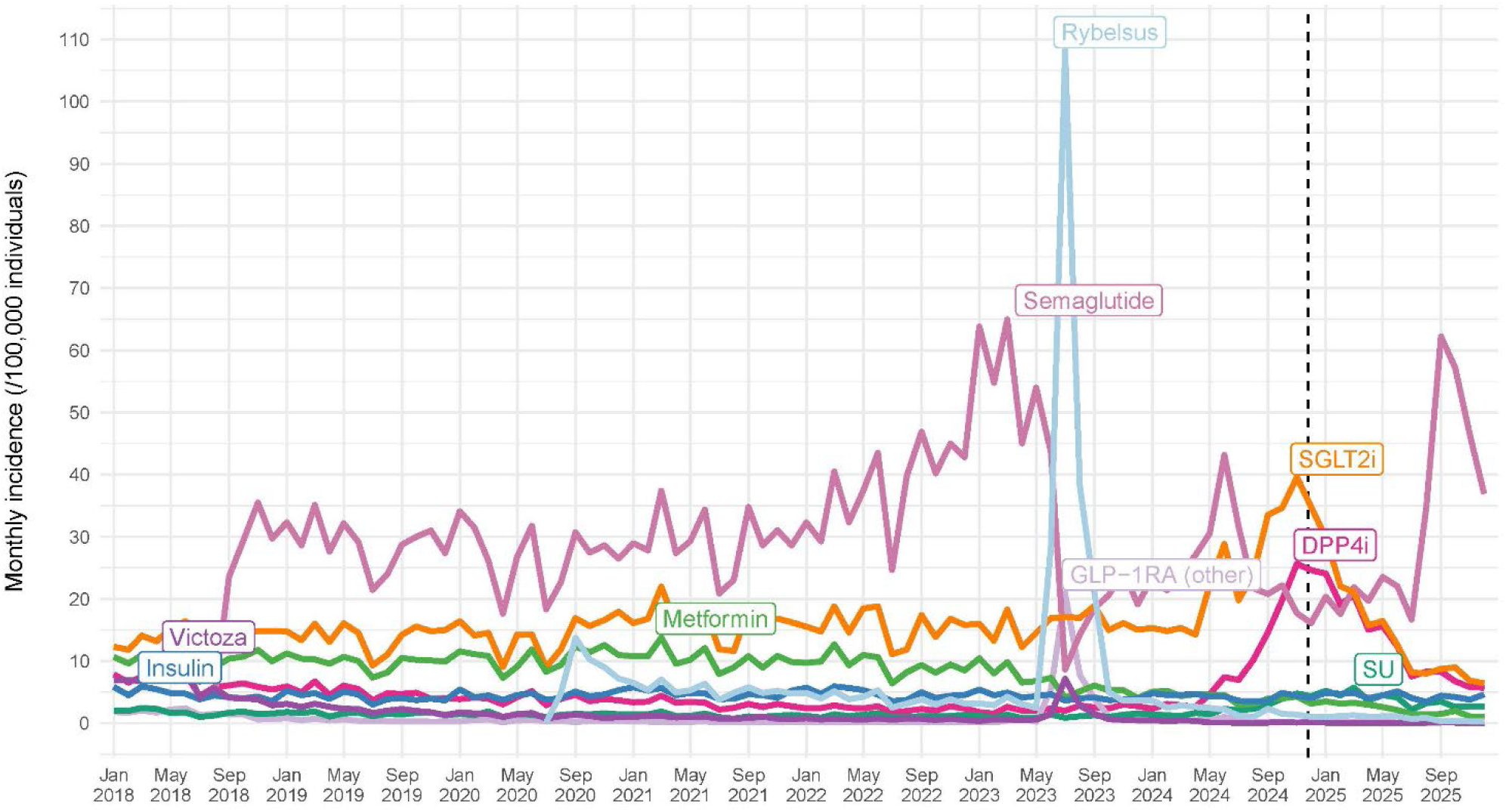
Monthly incidence rate of filled prescriptions for glucose-lowering drug classes in Denmark from January 2018 to December 2025. Trends are shown for semaglutide, Rybelsus®, Victoza®, other glucagon-like peptide-1 receptor agonists (GLP-1RA other), sodium-glucose cotransporter 2 inhibitors (SGLT2i), dipeptidyl peptidase-4 inhibitors (DPP4i), sulfonylureas (SU), metformin, and insulin. The dashed line indicates the policy change on 25 November 2024.

No clear increase in overall HbA1c testing intensity was observed around the implementation period (**Figure S2**).

### Characterization of reimbursement-consistent injectable semaglutide continuation

Among individuals who continued injectable semaglutide treatment after 25 November 2024, we classified continuation according to whether it was consistent with the first reimbursement criterion, defined as patients unable to use SGLT2is due to intolerance or severe renal impairment and with either (1) established cardiovascular or renal disease (regardless of glycemic control) or (2) multiple cardiovascular risk factors with inadequate glycemic control on metformin.

Among the 109,128 individuals who had filled at least one semaglutide prescription before the policy change, 61,120 also filled at least one semaglutide prescription within six months after the policy change and were classified as continuers (**Figure 1**). Among these, 44% (27,143 of 61,120 individuals) received concurrent SGLT2i treatment in the three months before the policy change, while 56% (33,977 of 61,120 individuals) did not receive concurrent SGLT2i treatment. Among those without concurrent SGLT2i treatment, 5.5% (1,862 of 33,977 individuals) met the predefined first criterion for reimbursement-consistent continuation. A total of 2,301 individuals had measurable markers of SGLT2i intolerance or severe renal impairment; of these, 1,854 had established cardiovascular or renal disease, while eight individuals met the criterion based on multiple cardiovascular risk factors and inadequate glycemic control on metformin.

Among those without concurrent SGLT2 inhibitor treatment, the remaining 95% (32,115 of 33,977 individuals) were classified as not meeting the operationalized first reimbursement criterion. Of these, 16,517 had previously been treated with an SGLT2i and may have had SGLT2i intolerance not captured by our registry-based definitions, potentially fulfilling the first reimbursement criterion (if they also had cardiorenal disease or multiple cardiovascular risk factors) or the second reimbursement criterion (if they had inadequate glycemic control despite prior use of all relevant oral glucose-lowering therapies). In a post hoc descriptive analysis, assuming previous SGLT2 inhibitor use as a proxy for intolerance or inability to continue treatment, the estimated proportion of individuals potentially meeting the reimbursement criteria increased from 5.5% to 41% (13,980/33,977).

Baseline characteristics of individuals who continued injectable semaglutide treatment, stratified according to consistency with the first reimbursement criterion, are presented in **Table S4**. Individuals with reimbursement-consistent continuation were generally older, more often female, and had a substantially higher burden of cardiovascular, renal, and diabetes-related comorbidities compared with those not meeting the first reimbursement criterion. Sensitivity analyses using alternative cutoffs for eGFR (e.g., <25 mL/min/1.73 m²) and HbA1c (e.g., >48 mmol/mol) did not meaningfully change the number of individuals with reimbursement-consistent continuation (data not shown).

### Switching patterns

Among the 22,096 individuals discontinuing semaglutide between March and November 2024, approximately 40% switched to at least one other glucose-lowering therapy within six months of follow-up, according to the prespecified switch definition. The most common subsequent therapies were SGLT2is (24%) and DPP4is (15%), although switching categories were not mutually exclusive. Switching to metformin (3.4%), insulin (2.3%), and SUs (2.0%) was less common, while switching to glitazones was negligible (**Table S5**). Switching patterns were similar across sex, with slightly higher switching proportions among males. Of the 22,096 individuals who discontinued semaglutide, 37% (8,284 of 22,096 individuals) reinitiated semaglutide after six months of discontinuation.

When stratifying by baseline regimen, switching from injectable semaglutide monotherapy was most common to SGLT2is (24%) and, to a lesser extent, DPP4i (8.5%). In non-insulin-containing regimens, switching generally favored SGLT2is – particularly among those on semaglutide + metformin (42%) and those on the registry-defined combination of semaglutide + DPP4i + metformin (37%; although this corresponded to only 75 individuals) – whereas individuals on semaglutide + SGLT2i + metformin more often switched to DPP4is (33%). In insulin-containing regimens, switching was primarily to SGLT2is and, less frequently, DPP4is.

When stratifying by injectable semaglutide treatment duration, switching to SGLT2is was more common among individuals with ≥1 year of use than <1 year (28% vs. 12%), while switching to other drug classes did not differ substantially by duration. By dose, switching to SGLT2is was less frequent at 0.25 mg (18%) than at 0.5 mg and 1 mg (both 25%).

Across HbA1c categories, switching to SGLT2is or DPP4is was the most common treatment change. SGLT2is were the most frequent switch among individuals with HbA1c <54 mmol/mol, whereas DPP4is were most common among those with HbA1c ≥54 mmol/mol. Switching to insulin became more frequent with increasing HbA1c, reaching 7.7% among patients with HbA1c 65–70 mmol/mol, while switching to SUs remained uncommon across all HbA1c categories (0.9–4.7%).

### Analysis of HbA1c changes

Among individuals who discontinued injectable semaglutide and had available HbA1c measurements, a total of 13,017 individuals were included. Mean HbA1c increased from 49 mmol/mol during treatment to 57 mmol/mol at the first eligible off-semaglutide treatment measurement, corresponding to a mean within-person change of 7.4 mmol/mol (95% CI 7.2–7.6) (**Table S6**). The increase was similar across sex, with mean changes of 7.6 mmol/mol (95% CI 7.3–7.9) among females and 7.3 mmol/mol (95% CI 7.0–7.6) among males.

In sensitivity analyses varying the definition of the off-semaglutide treatment HbA1c measurement and the handling of potential stockpiling, results were largely unchanged (**Table S6**). When using the latest off-treatment value, the mean change was 7.1 mmol/mol (95% CI 6.9–7.3). The median time from the last semaglutide fill to the latest off-treatment HbA1c measurement was 288 days (IQR 237–329).

When excluding potential stockpiling, the mean change was 7.0 mmol/mol (95% CI 6.8–7.3). Findings were similarly consistent across clinically relevant subgroups, including individuals switching to SGLT2is, those with shorter diabetes duration, and those with and without diabetes complications.

The change in HbA1c was related to baseline HbA1c: individuals in the lowest baseline categories had the largest increases – 8.1 mmol/mol (95% CI 7.9–8.3) in the ≤48 mmol/mol group and 9.5 mmol/mol (95% CI 9.0–10.0) in the 49–53 mmol/mol group – while those with baseline HbA1c ≥70 mmol/mol showed a mean decrease of 2.6 mmol/mol (95% CI -4.1 to -1.2).

Among 3,735 individuals meeting the SGLT2i-switch definition, 35% (1,324 of 3,735 individuals) re-initiated semaglutide within 12 months after switching to SGLT2is.

### Complementary analyses of HbA1c changes (interrupted time series)

Among individuals discontinuing injectable semaglutide, the estimated difference in monthly mean HbA1c between observed and counterfactual prediction levels was 5.9 mmol/mol (95% CI 5.1–6.7) at six months after treatment discontinuation, and 3.1 mmol/mol (95% CI 2.2–4.1) at 12 months. To assess longer-term HbA1c levels, we additionally examined measurements from 12 to 18 months after discontinuation; these measurements were not included in the analysis and showed relatively stable HbA1c levels that remained above pre-discontinuation levels (**Figure 4** and **Table S7**).

**Figure 4.**
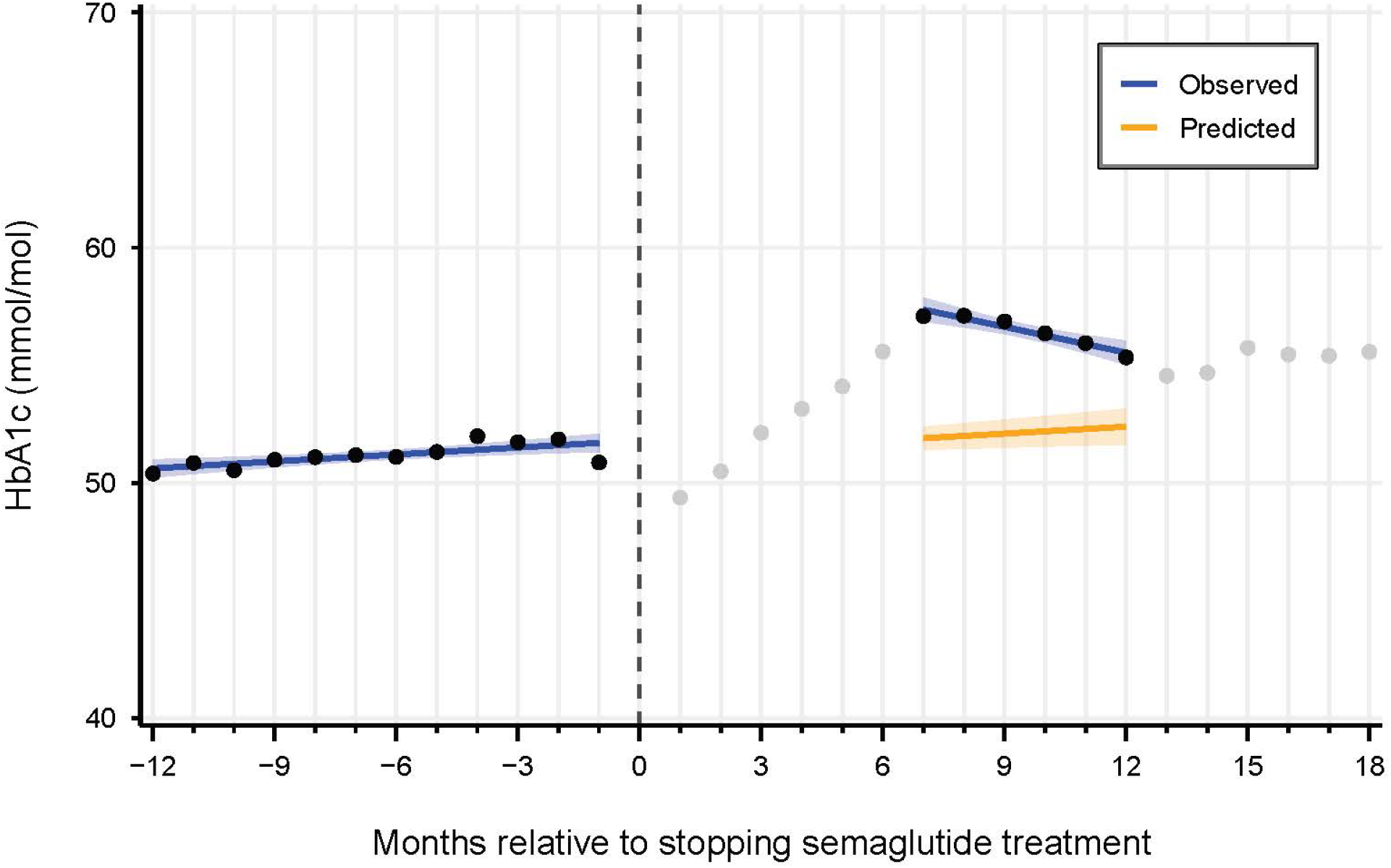
Interrupted time series of monthly mean HbA1c 12 months before and 6–12 months after semaglutide discontinuation (N = 21,264). The black points represent observed HbA1c values included in the analysis. The blue line shows the fitted linear model based on the observed data points. The orange line is the counterfactual prediction. The grey points represent observed HbA1c values not included in the analysis. Shaded areas indicate 95% confidence intervals.

Among individuals continuing injectable semaglutide treatment after 25 November 2024, the estimated difference in monthly mean HbA1c between observed and counterfactual prediction levels was 1.4 mmol/mol (95% CI 0.2–2.6) at six months after 25 November 2024, and 1.2 mmol/mol (95% CI -0.2 to 2.6) at 12 months. To assess longer-term HbA1c levels, we additionally examined measurements from 12 to 15 months after 25 November 2024, reflecting the available laboratory data through February 2026; these measurements were not included in the analysis and showed a slight increase in HbA1c levels (**Figure S3** and **Table S7**).

In a sensitivity analysis extending the post-policy assessment window to 3–12 months after the policy change, the observed monthly mean HbA1c values among individuals discontinuing injectable semaglutide were non-linear and were not adequately captured by the prespecified segmented linear regression model. Because the prespecified linear model did not adequately describe the observed pattern, we did not consider model-based estimates appropriate and therefore did not report them; **Figure S4** shows the observed monthly mean HbA1c values. Among individuals switching to SGLT2is, the estimated difference in monthly mean HbA1c between observed and counterfactual prediction levels was 8.7 mmol/mol (95% CI 7.6–9.8) at six months after treatment switching, and 3.8 mmol/mol (95% CI 2.5–5.1) at 12 months. To assess longer-term HbA1c levels, we additionally examined measurements from 12 to 18 months after treatment switching; these measurements were not included in the analyses and showed relatively stable HbA1c levels that remained above pre-switch levels (**Figure S5** and **Table S7**).

## Discussion

In this nationwide registry-based study, initiation of injectable semaglutide for diabetes appeared to decline around the time of the policy change, followed by a subsequent increase in 2025, consistent with national data showing a rebound in injectable semaglutide use later that year.^23^ Among individuals with a semaglutide fill between March and November 2024, nearly three-quarters continued treatment through 25 November 2024. Among continuers without concurrent SGLT2i use, only a small minority fulfilled the first reimbursement criterion (non-feasibility of SGLT2i use and high cardiovascular risk), based on the information available in the registries. Among individuals who discontinued semaglutide, most did not appear to initiate another glucose-lowering therapy during six months of follow-up. At the same time, HbA1c increased after discontinuation and remained above pre-discontinuation levels during 18 months of follow-up.

This study has several important strengths. Danish nationwide health registries provide near-complete population coverage, minimizing selection bias and loss to follow-up. Linkage of prescription, hospital, and laboratory data enabled detailed characterization of treatment patterns before and after the reimbursement change. Access to HbA1c measurements allowed assessment of short-term clinical consequences of semaglutide discontinuation. However, several limitations should be considered. Full reimbursement eligibility could not be assessed, as the available data allowed operationalization of only one of the two criteria. Also, prescription fills do not necessarily reflect actual medication use, and despite accounting for stockpiling and irregular refills, discontinuation timing may still have been imprecise, potentially weakening observed HbA1c changes. Individuals continuing semaglutide after the policy change likely reflected both reimbursed and self-funded treatment, which could not be distinguished. Our assessment of treatment switching focused on initiation of new glucose-lowering therapies and did not capture dose escalation of existing treatments. Finally, HbA1c analyses were restricted to individuals with available laboratory measurements and may therefore reflect a selected subgroup under closer clinical monitoring.

The finding that only 5.5% of individuals fulfilled the operationalized reimbursement criterion should be interpreted with caution, as only one of the two reimbursement criteria could be operationalized using registry data. The second criterion relied on individualized clinical assessment, including prior use of all “relevant” oral glucose-lowering therapies and patient-specific HbA1c targets, which were unavailable. Other factors influencing treatment decisions, including tolerability, symptom burden, patient preferences, body mass index, and clinical judgment, were also not captured. In a supplementary analysis using previous SGLT2i use as a proxy for intolerance, 41% fulfilled the operationalized criterion. However, previous SGLT2i use does not establish intolerance and may overestimate eligibility, whereas the primary estimate of 5.5% may underestimate it. Together, these analyses provide important context for interpreting the implementation of the revised reimbursement criteria.

Most individuals who discontinued semaglutide did not switch to another glucose-lowering therapy, suggesting that discontinuation was generally not followed by substitution with another glucose-lowering drug. Notably, nearly one out of three semaglutide discontinuers restarted their semaglutide therapy more than six months after discontinuation. Among those who did switch, the observed patterns appeared broadly consistent with contemporary T2D treatment recommendations. Switching to SGLT2is predominated, likely reflecting prioritization of drugs with established cardiovascular and renal benefits, particularly among patients at elevated cardiovascular risk.^4–7^ Switching to DPP4is may represent treatment choices in lower-risk populations where multiple alternatives are clinically acceptable.^3^ Switching to SUs was uncommon, consistent with caution regarding hypoglycemia risk, particularly among older individuals and patients with comorbidities.^3^ Few individuals switched to metformin, likely because many were already receiving it or could not tolerate it.^3^ Overall, the switching patterns suggest that treatment changes following semaglutide discontinuation were heterogeneous and likely reflected individual treatment considerations.

Discontinuation of injectable semaglutide was followed by an initial increase in HbA1c. Although this increase subsequently declined, levels remained modestly above pre-discontinuation levels through 18 months of follow-up. Increases were largest among patients with lower baseline HbA1c, whereas those with baseline HbA1c ≥70 mmol/mol experienced a decrease – a plausible pattern given that well-controlled patients typically have more room to drift upward before clinical intervention follows; this post-hoc finding moderates the clinical implications of the reported worsened glycemic control. While downstream clinical outcomes were not assessed, long-term cohort data showed that as people’s running-average HbA1c rises, their complication risk rises steadily and continuously – with no flat zone and no sudden jumps along the way.^24^ This suggests that sustained increases of this magnitude are unlikely to be risk-neutral, even though their translation to hard outcomes over a period this short remains unclear. An increase of this size is expected following discontinuation of an effective glucose-lowering therapy and is not necessarily specific to semaglutide. Although some discontinuations occurred before the winter and holiday season, seasonal variation alone is unlikely to explain the findings, as no comparable increase was observed among semaglutide continuers.

The implications of our findings for policymakers are complex. Previous studies show that reimbursement changes can shift glucose-lowering treatment and that increases in medication costs may worsen glycemic control.^25,26^ Our findings extend this evidence to a national restriction targeting a modern GLP-1RA, showing that discontinuation was often not followed by treatment substitution and was associated with an increase in HbA1c that remained above pre-discontinuation levels during 18 months of follow-up. Similar deterioration has been observed following GLP-1RA shortages.^27,28^ Although reimbursement restrictions may promote more efficient use, their implementation should account for appropriate treatment modification and follow-up of glycemic control among patients who lose access to established therapy.

In conclusion, the November 2024 reimbursement policy change on GLP-1RA prescriptions in Denmark affected treatment patterns and short-term glycemic control among individuals with T2D treated with semaglutide. Treatment switching occurred predominantly toward SGLT2is, while semaglutide discontinuation was associated with an increase in HbA1c that remained above pre-discontinuation levels during follow-up.

## Data Availability

The lead author (AP) affirms that the manuscript is an honest, accurate, and transparent account of the study being reported; that no important aspects of the study have been omitted; and that any discrepancies from the study as originally planned have been explained. The study protocol was registered at the Real-World Evidence Registry (https://osf.io/jbzda) before the commencement of any analyses, with subsequent amendments also made available (https://osf.io/gxhp3). Due to Danish data protection regulations, individual-level data cannot be shared directly by the authors. The source code used for data management and statistical analyses is available at https://gitlab.sdu.dk/pharmacoepi/Ozempic-Reimbursement.

## Declarations

## Acknowledgements

Jacob Harbo Andersen from the University of Southern Denmark is acknowledged for his validation of the study code. He was not otherwise involved in the study.

## Approvals

The study was registered at the repository of the University of Southern Denmark (12.745), and data access was granted by the Danish Health Data Authority (FSEID-6047). Ethical approval is not required in Denmark for purely registry-based studies.

## Data sharing

Data were used under license and thus cannot be made publicly available. De-identified data can be made available for authorized researchers after an application to ForskerService at the Danish Health Data Authority. The source code used to perform data management and statistical analyses is available at https://gitlab.sdu.dk/pharmacoepi/Ozempic-Reimbursement.

## Author contributions (including data access and responsibility)

The study was designed by authors ES and AP. MTE and AP had access to the Danish data, and they take responsibility for the integrity and accuracy of their data and analyses. The first draft of the manuscript was written by ES and AP and revised for important intellectual content by all authors. The final version of the paper was approved by all authors.

## SUPPLEMENTARY

### Supplemental methods

#### Analysis of HbA1c changes (paired t-test)

We performed predefined subgroup analyses for the analysis of HbA1c changes according to sex, age group (<40, 40–59, 60–79, and ≥80 years), baseline HbA1c (≤48, 49–53, 54–58, 59–64, 65–70, and ≥70 mmol/mol), diabetes duration (<5 and ≥5 years), and the presence or absence of diabetes-related complications, including cardiovascular, renal, neurological, and ocular complications.

To assess the potential impact of stockpiling before treatment discontinuation, we conducted a sensitivity analysis excluding individuals with potential stockpiling, defined based on the timing and quantity of their final semaglutide fill. Specifically, we excluded individuals who either redeemed their final prescription within one month before the policy change or had an unusually large final fill, defined as more than 90 defined daily doses and more than twice the quantity dispensed at the previous fill.

As a complementary analysis, we examined patients switching to SGLT2is, defined as the first SGLT2i prescription within three months after the last semaglutide fill, with no SGLT2i fills in the preceding two years. HbA1c values before and after the switch were compared using the same within-individual approach as in the primary analysis. We additionally described subsequent reinitiation of semaglutide among these individuals, including the frequency and timing of reinitiations.

---

**Table S1.** ATC codes used to define drug use any time before the index date.

|  |  |
| --- | --- |
| Ozempic® (semaglutide) | ATC: A10BJ06 with drug ids<br>032822 046216 063436 072557 081586 089980 099045 110341 126158 144807<br>150408 178275 183356 183727 185998 195994 408233 417633 417977 435625<br>500302 502065 504236 505084 545670 582611 586531 592316 |
| Rybelsus® (semaglutide) | ATC: A10BJ06 with drug ids<br>063666 127918 135230 150218 154023 158785 185980 377782 415557 424430<br>432723 435469 452499 455632 466316 471211 478638 485999 534525 540676<br>565251 578896 |
| Victoza® (liraglutide) | ATC: A10BJ02 with drug ids<br>052838 058837 071445 100378 125371 375726 379220 398806 435653 449008<br>451425 457211 483901 492860 504075 592557 597455 |
| ‘Other’ GLP-1RAs | A10BJ01 (exenatide), A10BJ03 (lixisenatide), A10BJ05 (dulaglutide), A10AE54 (insulin glargine and lixisenatide), A10AE56 (insulin degludec and liraglutide) |
| SGLT2is | A10BK*<br>A10BD[15,16,19,20,21,23,24,25,27,29,30,31,32,33,34] (combinations) |
| DPP4is | A10BH*<br>A10BD[07,08,09,10,11,12,13,18,19,21,22,24,25,27,28,29,30,31] (combinations) |
| SUs | A10BB*<br>A10BD[01,02,04,06] (combinations)<br>A10BC01 |
| Metformin | A10BA*<br>A10BD[02,03,05,07,08,10,11,13,14,15,16,17,18,20,22,23,25,26,27,28,31,34] (combinations) |
| Insulins | A10A* |
| Glitazones | A10BG* |
| Statins | C10AA*<br>C10BA* (combinations)<br>C10BX* (combinations) |
| Thromboprophylaxis | B01AC06, B01AC04, B01AC22, B01AC07, B01AC24, B01AC25 |
| Psychoactive medications | N05A, N05BA, N05CD, N05CF, N06A |
| Abbreviations: GLP-1RAs; glucagon-like peptide-1 receptor agonists; SGLT2is; sodium-glucose cotransporter 2 inhibitors; DPP4is; dipeptidyl peptidase-4 inhibitors; SUs, sulfonylureas. |  |

**Table S2.** Markers of diabetes severity and comorbidity. The index date for discontinuers was defined as the date of their last Ozempic fill. The index date for continuers was the date of the policy change (i.e., 25 November 2024). Unless otherwise specified in the Methods section, these index dates and corresponding time windows were used throughout the analyses.

| Variable | Coding system | Window | Codes |
| --- | --- | --- | --- |
| HbA1c, mmol/mol | NPU | [-365 days; -1 day]<br>[+182 days; +365 days] | NPU03835, NPU27300 |
| eGFR (estimated based on creatinine values using the CKD-EPI formula) | NPU | Most recent value within two years before the index date. | NPU01807, NPU04998, NPU09101, NPU17559, NPU18016, NPU18105 |
| uACR | NPU | Any time before the index date. | NPU19661, NPU03918 |
| Ischemic heart disease | ICD-10 | Any time before the index date. | I20, I21, I22, I23, I24, I25, T822, T823 |
| Cerebrovascular disease | ICD-10 | Any time before the index date. | I60, I61, I63, I64, I65, I66, I672, I678, I679, I691, I693, I694, I695, I696, I697, I698 |
| Atherosclerotic peripheral vascular disease | ICD-10 | Any time before the index date. | E105, E115, E125, E135, E145, I70, I739, I74, K550, K551, N280 |
| Heart failure | ICD-10 | Any time before the index date. | I50, I110, I130, I132, I420, I426, I427, I428, I429 |
| Renal complications | ICD-10 | Any time before the index date. | E102, E112, E122, E132, E142, I12, I13, N01, N02, N03, N04, N05, N06, N083, N17, N18, N19, R809, Z992 |
|  | SKS | Any time before the index date. | BJFD0, BJFD2, KKAS00, KKAS10, KKAS20 |
|  | Laboratory values | Any time before the index date. | eGFR <60 mL/min/1.73 m<sup>2</sup>† and/or uACR ≥30 mg/g, measured on at least two occasions with a minimum interval of three months between measurements |
| Neurological complications | ICD-10 | Any time before the index date. | E104, E114, E124, E134, E144, G590, G598, G603, G609, G618, G619, G620, G621, G622, G628, G629, G630, G631, G632, G634, G635, G636, G638, G990 |
| Ocular complications | ICD-10 | Any time before the index date. | E103, E113, E123, E133, E143, H280, H330, H332, H334, H335, H340, H341, H342, H360, H430, H431, H438C, H439, H450, H470, H540, H541, H544 |
|  | SKS | Any time before the index date. | KCKC10, KCKC15, KCKD65 |
| Foot ulcers | ICD-10 | Any time before the index date. | E105B-E145B, E105C-E145C, L97, R02, S91, L024A, L024J, L024M |
| Hypertension | ICD-10 | Within two years before the index date. | I10, I11, I12, I13, I14, I15 |
|  | ATC | Within two years before the index date. | C02A, C02B, C02C (α-adrenergic agents)<br>C03A, C03EA (thiazides)<br>C03D, C03E (potassium-sparing agents)<br>C07 (beta-blockers)<br>C08 (calcium channel antagonists)<br>C09 (renin-angiotensin-system antagonists) |
| Smoking-related disorders | ICD-10 | Within two years before the index date. | J40, J41, J42, J43, J44, J47, J961, Z587, Z720 |
|  | ATC | Within two years before the index date. | N06AX12, N07BA |
| Alcohol abuse | ICD-10 | Any time before the index date. | E244, E529, F10, G312, G621, G721, I85, I426, I864, I982, K70, K292, K852, K860, L278A, T51, Z714, Z721 |
|  | ATC | Any time before the index date. | N07BB01, N07BB03 |
| Obesity | ICD-10 | Within two years before the index date. | E66, E68 |
| Chronic liver disease | ICD-10 | Any time before the index date. | B16, B18, B150, B190, I85, K70, K71, K72, K73, K74, K760, K766 |
| Diabetic ketoacidosis | ICD-10 | Any time before the index date. | E101, E111, E121, E131, E141 |
| Hypoglycemia | ICD-10 | Any time before the index date. | E100, E110, E120, E130, E140, E159, E160, E161, E162 |
| Pancreatitis | ICD-10 | Any time before the index date. | K85, K860, K861 |
| Pancreas cancer | ICD-10 | Any time before the index date. | C25 |
| Thyroid cancer | ICD-10 | Any time before the index date. | C73 |
| UTI | ICD-10 | Within two years before the index date. | N10, N11, N12, N136, N151, DN30 except N304, N340, N341, N390 |
|  | ATC | Within two years before the index date. | J01CA08, J01EA, J01EB02, J01XE01 |
| GTI | ICD-10 | Within two years before the index date. | B373, B374, N481, N493, N771, N760, N761, N762, N763, N7682 |
|  | ATC | Within the year before the index date. | D01AC01 |
| Dyslipidemia | ICD-10 | Within two years before the index date. | E78 |
|  | ATC | Within two years before the index date. | C10AA*<br>C10BA* (combinations)<br>C10BX* (combinations) |
| LDL cholesterol | NPU | Most recent value within two years before the index date. | NPU01568, NPU10171 |
| HDL cholesterol | NPU | Most recent value within two years before the index date. | NPU01567, NPU10157, NPU18107 |
| Triglycerides | NPU | Most recent value within two years before the index date. | NPU03620, NPU04094, NPU18106 |
| Total cholesterol | NPU | Most recent value within two years before the index date. | NPU01566 |
| Abbreviations: NPU, Nomenclature for Properties and Units; ICD-10, International Classification of Diseases, Tenth Revision; SKS, The Danish Medical Classification System; eGFR, estimated glomerular filtration rate; uACR, urine albumin-creatinine ratio; UTI, urinary tract infection; GTI, genital tract infection; LDL, low-density lipoprotein; HDL, high-density lipoprotein.<br>†Based on the most recent outpatient or primary care eGFR before the index date (inpatient measurements excluded). |  |  |  |

**Table S3.** Baseline characteristics of semaglutide users who discontinued treatment between March and November 2024 versus those who continued treatment through 25 November 2024.

|  | **Continuation of semaglutide**<br>**(N = 57,209)** | **Discontinuation of**<br>**semaglutide (N = 22,096)** |
| --- | --- | --- |
| Female sex, N (%) | 24,450 (43) | 10,740 (49) |
| Age, years, median (IQR) | 61 [53, 69] | 60 [51, 69] |
| Hypertension | 47,191 (82) | 16,757 (76) |
| Dyslipidemia | 47,172 (82) | 16,517 (75) |
| Ocular disease | 9,339 (16) | 2,470 (11) |
| Renal disease | 30,488 (53) | 10,518 (48) |
| GTI | 565 (1.0) | 158 (0.7) |
| UTI | 9,811 (17) | 3,777 (17) |
| Markers of smoking | 1,804 (3.2) | 755 (3.4) |
| Markers of alcohol abuse | 3,984 (7.0) | 1,684 (7.6) |
| PVD | 6,523 (11) | 1,859 (8.4) |
| Pancreas cancer | 68 (0.1) | 24 (0.1) |
| Thyroid cancer | 179 (0.3) | 78 (0.4) |
| Cerebrovascular disease | 5,289 (9.2) | 1,802 (8.2) |
| Heart failure | 4,639 (8.1) | 1,276 (5.8) |
| Hypoglycemia | 1,470 (2.6) | 460 (2.1) |
| Ischemic heart disease | 12,774 (22) | 3,739 (17) |
| Diabetic ketoacidosis | 829 (1.4) | 240 (1.1) |
| Chronic liver disease | 2,518 (4.4) | 980 (4.4) |
| Neurological disease | 7,379 (13) | 1,889 (8.5) |
| Obesity | 3,109 (5.4) | 1,218 (5.5) |
| Pancreatitis | 1,308 (2.3) | 483 (2.2) |
| Psychoactive medications | 34,456 (60) | 13,381 (61) |
| Statins | 50,542 (88) | 17,831 (81) |
| Thromboprophylaxis | 27,029 (47) | 8,436 (38) |
| N (%) with a lab value for uACR | 49,929 (87) | 17,970 (81) |
| uACR, median (IQR) | 14 [7.0, 38] | 12 [6.0, 31] |
| N (%) with a lab value for eGFR | 56,862 (99) | 21,828 (99) |
| eGFR, median (IQR) | 74 [62, 91] | 72 [61, 87] |
| N (%) with a lab value for HbA1c | 56,708 (99) | 21,727 (98) |
| HbA1c, median (IQR) | 51 [44, 59] | 46 [40, 55] |
| N (%) with a lab value for LDL cholesterol | 35,112 (61) | 13,206 (60) |
| LDL cholesterol, median (IQR) | 1.7 [1.3, 2.3] | 1.8 [1.4, 2.4] |
| N (%) with a lab value for HDL cholesterol | 56,220 (98) | 21,455 (97) |
| HDL cholesterol, median (IQR) | 1.1 [0.9, 1.3] | 1.2 [1.0, 1.4] |
| N (%) with a lab value for triglycerides | 56,403 (99) | 21,536 (98) |
| Triglycerides, median (IQR) | 1.7 [1.2, 2.5] | 1.6 [1.2, 2.3] |
| N (%) with a lab value for total cholesterol | 56,271 (98) | 21,488 (97) |
| Total cholesterol, median (IQR) | 3.7 [3.2, 4.4] | 3.8 [3.3, 4.6] |
| DPP4i* | 24,326 (42) | 5,943 (28) |
| Other GLP-1RA* | 4,171 (7.3) | 943 (4.5) |
| SGLT2i* | 41,831 (73) | 9,909 (47) |
| SU* | 19,344 (34) | 4,820 (23) |
| Metformin* | 54,797 (96) | 19,774 (93) |
| Insulin* | 22,149 (39) | 5,623 (27) |
| Glitazone* | 843 (1.5) | 200 (0.9) |
| Rybelsus®* | 9,971 (17) | 2,983 (14) |
| Victoza®* | 18,256 (32) | 4,477 (21) |
| Abbreviations: IQR, interquartile range; GTI, genital tract infection; UTI, urinary tract infection; PVD, peripheral vascular disease; uACR, urine albumin-creatinine ratio; SD, standard deviation; eGFR, estimated glomerular filtration rate; LDL, low-density lipoprotein; HDL, high-density lipoprotein; DPP4i, dipeptidyl peptidase-4 inhibitor; GLP-1RA, glucagon-like peptide-1 receptor agonist; SGLT2i, sodium-glucose cotransporter 2 inhibitor; SU, sulfonylurea. *Use any time before the index date. |  |  |

**Table S4.**
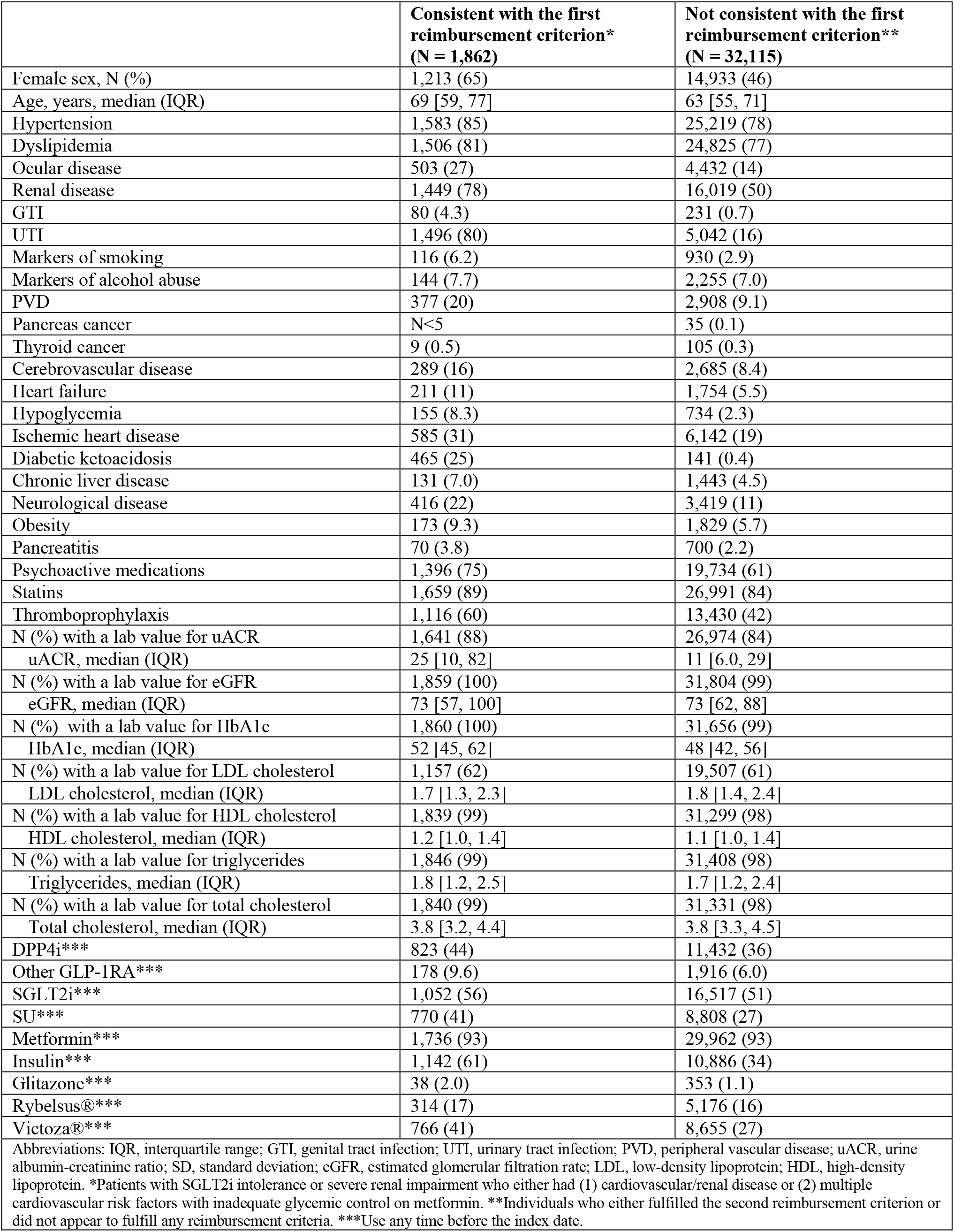
Baseline characteristics of semaglutide continuers, i.e., individuals with at least one semaglutide prescription before 25 November 2024 and at least one prescription within six months after 25 November 2024, according to the first reimbursement criterion*.

|  | **Consistent with the first reimbursement criterion\***<br>**(N = 1,862)** | **Not consistent with the first reimbursement criterion\*\***<br>**(N = 32,115)** |
| --- | --- | --- |
| Female sex, N (%) | 1,213 (65) | 14,933 (46) |
| Age, years, median (IQR) | 69 [59, 77] | 63 [55, 71] |
| Hypertension | 1,583 (85) | 25,219 (78) |
| Dyslipidemia | 1,506 (81) | 24,825 (77) |
| Ocular disease | 503 (27) | 4,432 (14) |
| Renal disease | 1,449 (78) | 16,019 (50) |
| GTI | 80 (4.3) | 231 (0.7) |
| UTI | 1,496 (80) | 5,042 (16) |
| Markers of smoking | 116 (6.2) | 930 (2.9) |
| Markers of alcohol abuse | 144 (7.7) | 2,255 (7.0) |
| PVD | 377 (20) | 2,908 (9.1) |
| Pancreas cancer | N<5 | 35 (0.1) |
| Thyroid cancer | 9 (0.5) | 105 (0.3) |
| Cerebrovascular disease | 289 (16) | 2,685 (8.4) |
| Heart failure | 211 (11) | 1,754 (5.5) |
| Hypoglycemia | 155 (8.3) | 734 (2.3) |
| Ischemic heart disease | 585 (31) | 6,142 (19) |
| Diabetic ketoacidosis | 465 (25) | 141 (0.4) |
| Chronic liver disease | 131 (7.0) | 1,443 (4.5) |
| Neurological disease | 416 (22) | 3,419 (11) |
| Obesity | 173 (9.3) | 1,829 (5.7) |
| Pancreatitis | 70 (3.8) | 700 (2.2) |
| Psychoactive medications | 1,396 (75) | 19,734 (61) |
| Statins | 1,659 (89) | 26,991 (84) |
| Thromboprophylaxis | 1,116 (60) | 13,430 (42) |
| N (%) with a lab value for uACR | 1,641 (88) | 26,974 (84) |
| uACR, median (IQR) | 25 [10, 82] | 11 [6.0, 29] |
| N (%) with a lab value for eGFR | 1,859 (100) | 31,804 (99) |
| eGFR, median (IQR) | 73 [57, 100] | 73 [62, 88] |
| N (%) with a lab value for HbA1c | 1,860 (100) | 31,656 (99) |
| HbA1c, median (IQR) | 52 [45, 62] | 48 [42, 56] |
| N (%) with a lab value for LDL cholesterol | 1,157 (62) | 19,507 (61) |
| LDL cholesterol, median (IQR) | 1.7 [1.3, 2.3] | 1.8 [1.4, 2.4] |
| N (%) with a lab value for HDL cholesterol | 1,839 (99) | 31,299 (98) |
| HDL cholesterol, median (IQR) | 1.2 [1.0, 1.4] | 1.1 [1.0, 1.4] |
| N (%) with a lab value for triglycerides | 1,846 (99) | 31,408 (98) |
| Triglycerides, median (IQR) | 1.8 [1.2, 2.5] | 1.7 [1.2, 2.4] |
| N (%) with a lab value for total cholesterol | 1,840 (99) | 31,331 (98) |
| Total cholesterol, median (IQR) | 3.8 [3.2, 4.4] | 3.8 [3.3, 4.5] |
| DPP4i*** | 823 (44) | 11,432 (36) |
| Other GLP-1RA*** | 178 (9.6) | 1,916 (6.0) |
| SGLT2i*** | 1,052 (56) | 16,517 (51) |
| SU*** | 770 (41) | 8,808 (27) |
| Metformin*** | 1,736 (93) | 29,962 (93) |
| Insulin*** | 1,142 (61) | 10,886 (34) |
| Glitazone*** | 38 (2.0) | 353 (1.1) |
| Rybelsus®*** | 314 (17) | 5,176 (16) |
| Victoza®*** | 766 (41) | 8,655 (27) |
| Abbreviations: IQR, interquartile range; GTI, genital tract infection; UTI, urinary tract infection; PVD, peripheral vascular disease; uACR, urine albumin-creatinine ratio; SD, standard deviation; eGFR, estimated glomerular filtration rate; LDL, low-density lipoprotein; HDL, high-density lipoprotein. *Patients with SGLT2i intolerance or severe renal impairment who either had (1) cardiovascular/renal disease or (2) multiple cardiovascular risk factors with inadequate glycemic control on metformin. **Individuals who either fulfilled the second reimbursement criterion or did not appear to fulfill any reimbursement criteria. ***Use any time before the index date. |  |  |

**Table S5.** Switching to alternative glucose-lowering therapies among individuals discontinuing semaglutide between March and November 2024. Percentages are shown overall and stratified by sex, baseline glucose-lowering regimen, duration of semaglutide use, last semaglutide dose, and baseline HbA1c levels. Percentages do not sum to 100% because individuals could switch to more than one glucose-lowering drug during follow-up.

|  | N | Did not switch to alternative drug within 6 months*, n (%) | Percentage of patients switching to alternative drug**, n (%) |  |  |  |  |  |
| --- | --- | --- | --- | --- | --- | --- | --- | --- |
|  |  |  | SGLT2i | DPP4i | SU | Metformin | Insulin | Glitazone |
| **Overall** | 22,096 | 13,216 (60%) | 5,300 (24%) | 3,227 (15%) | 446 (2.0%) | 750 (3.4%) | 505 (2.3%) | 0 (0.0%) |
| **Female** | 10,740 | 6,578 (61%) | 2,517 (23%) | 1,502 (14%) | 205 (1.9%) | 380 (3.5%) | 243 (2.3%) | 0 (0.0%) |
| **Male** | 11,356 | 6,638 (58%) | 2,783 (25%) | 1,725 (15%) | 241 (2.1%) | 370 (3.3%) | 262 (2.3%) | 0 (0.0%) |
| **Baseline regimen (incl. semaglutide)** |  |  |  |  |  |  |  |  |
| Monotherapy semaglutide | 6,642 | 4,254 (64%) | 1,602 (24%) | 567 (8.5%) | 75 (1.1%) | 577 (8.7%) | 95 (1.4%) | 0 (0.0%) |
| SGLT2i | 1,188 | 754 (63%) | 0 (0.0%) | 309 (26%) | 36 (3.0%) | 90 (7.6%) | 54 (4.5%) | 0 (0.0%) |
| Metformin | 6,961 | 3,346 (48%) | 2,956 (42%) | 972 (14%) | 116 (1.7%) | 0 (0.0%) | 118 (1.7%) | 0 (0.0%) |
| Insulin | 1,057 | 788 (75%) | 188 (18%) | 61 (5.8%) | <5 | 47 (4.4%) | 0 (0.0%) | 0 (0.0%) |
| SGLT2i + Metformin | 3,015 | 1,805 (60%) | 0 (0.0%) | 993 (33%) | 172 (5.7%) | 0 (0.0%) | 146 (4.8%) | 0 (0.0%) |
| SGLT2i + Insulin | 418 | 354 (85%) | 0 (0.0%) | 48 (11%) | <5 | 18 (4.3%) | 0 (0.0%) | 0 (0.0%) |
| DPP4i + Metformin*** | 204 | 106 (52%) | 75 (37%) | 0 (0.0%) | 11 (5.4%) | 0 (0.0%) | 17 (8.3%) | 0 (0.0%) |
| Metformin + Insulin | 1,043 | 610 (58%) | 379 (36%) | 90 (8.6%) | 6 (0.6%) | 0 (0.0%) | 0 (0.0%) | 0 (0.0%) |
| SGLT2i + Metformin + Insulin | 823 | 711 (86%) | 0 (0.0%) | 110 (13%) | <5 | 0 (0.0%) | 0 (0.0%) | 0 (0.0%) |
| Other insulin-containing regimens | 123 | 93 (76%) | 22 (18%) | 9 (7.3%) | <5 | <5 | 0 (0.0%) | 0 (0.0%) |
| Other metformin-containing regimens | 419 | 267 (64%) | 49 (12%) | 51 (12%) | 16 (3.8%) | 0 (0.0%) | 49 (12%) | 0 (0.0%) |
| Other treatment regimens | 203 | 128 (63%) | 29 (14%) | 17 (8.4%) | <5 | >13 | 26 (13%) | 0 (0.0%) |
| **Duration of semaglutide treatment** |  |  |  |  |  |  |  |  |
| <1 year | 5,525 | 4,089 (74%) | 652 (12%) | 593 (11%) | 103 (1.9%) | 107 (1.9%) | 145 (2.6%) | 0 (0.0%) |
| ≥1 year | 16,571 | 9,127 (55%) | 4,648 (28%) | 2,634 (16%) | 343 (2.1%) | 643 (3.9%) | 360 (2.2%) | 0 (0.0%) |
| **Last semaglutide dose** |  |  |  |  |  |  |  |  |
| 0.25 mg | 2,868 | 1,856 (65%) | 512 (18%) | 366 (13%) | 61 (2.1%) | 82 (2.9%) | 96 (3.3%) | 0 (0.0%) |
| 0.5 mg | 6,814 | 3,963 (58%) | 1,716 (25%) | 1,042 (15%) | 129 (1.9%) | 216 (3.2%) | 150 (2.2%) | 0 (0.0%) |
| 1 mg | 12,414 | 7,397 (60%) | 3,072 (25%) | 1,819 (15%) | 256 (2.1%) | 452 (3.6%) | 259 (2.1%) | 0 (0.0%) |
| **HbA1c levels** |  |  |  |  |  |  |  |  |
| ≤48 mmol/mol | 9,917 | 6,114 (62%) | 2,622 (26%) | 1,158 (12%) | 89 (0.9%) | 370 (3.7%) | 61 (0.6%) | 0 (0.0%) |
| 49–53 mmol/mol | 2,538 | 1,224 (48%) | 719 (28%) | 611 (24%) | 93 (3.7%) | 82 (3.2%) | 45 (1.8%) | 0 (0.0%) |
| 54–58 mmol/mol | 1,663 | 919 (55%) | 344 (21%) | 374 (22%) | 74 (4.4%) | 32 (1.9%) | 56 (3.4%) | 0 (0.0%) |
| 59–64 mmol/mol | 1,215 | 680 (56%) | 232 (19%) | 246 (20%) | 57 (4.7%) | 26 (2.1%) | 63 (5.2%) | 0 (0.0%) |
| 65–70 mmol/mol | 750 | 478 (64%) | 102 (14%) | 124 (17%) | 20 (2.7%) | 25 (3.3%) | 58 (7.7%) | 0 (0.0%) |
| Abbreviations: SU, sulfonylurea; DPP4i, dipeptidyl peptidase-4 inhibitor; SGLT2i, sodium-glucose cotransporter 2 inhibitor. *No switching within six months of discontinuing semaglutide for diabetes management. **Switching categories were not mutually exclusive, i.e., switching to more than one drug class was possible. ***Registry-defined combination, as concomitant use of DPP4i and semaglutide is generally not prescribed in clinical practice because it provides no meaningful additional glycemic benefit. |  |  |  |  |  |  |  |  |

**Table S6.**
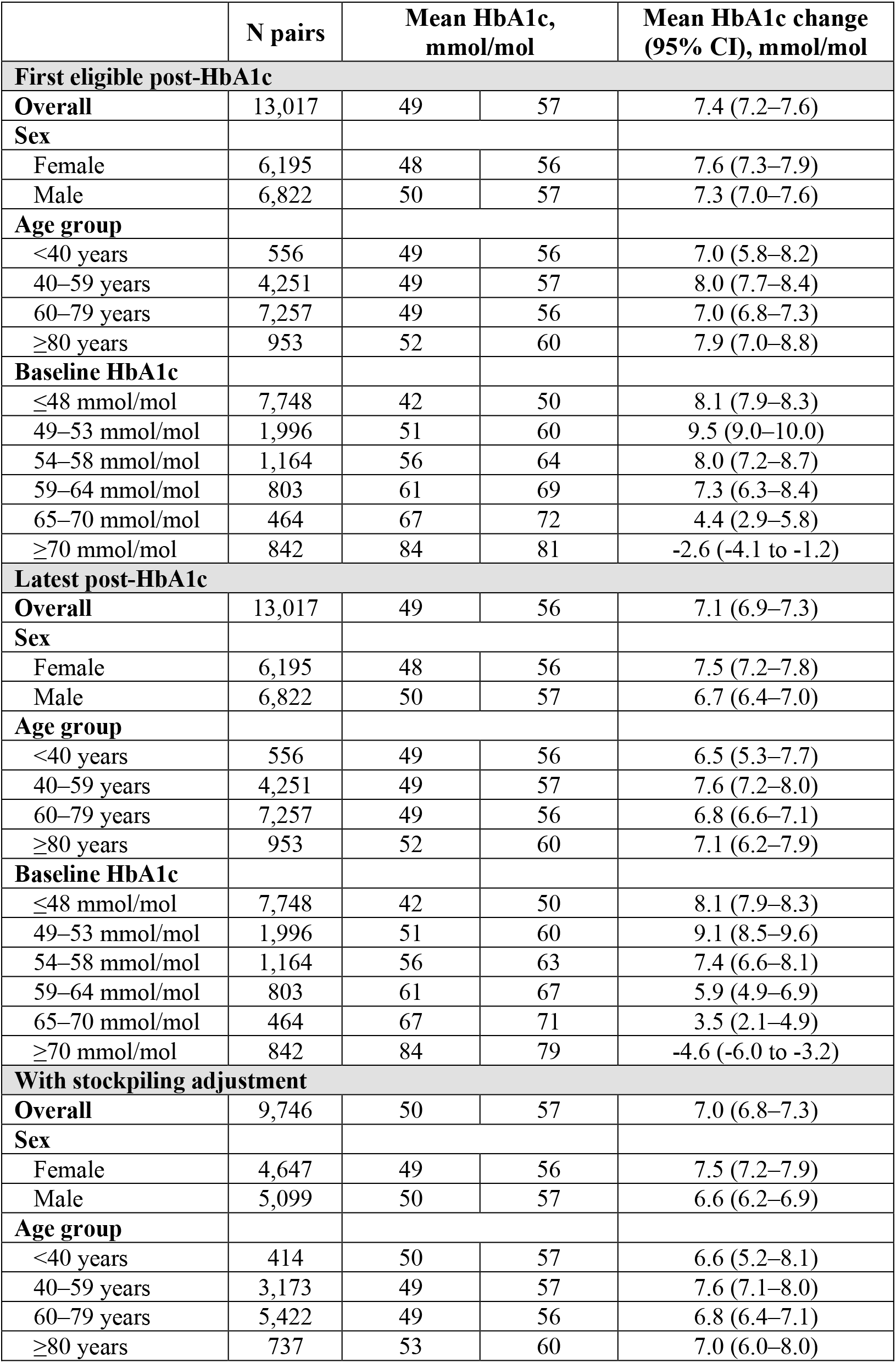
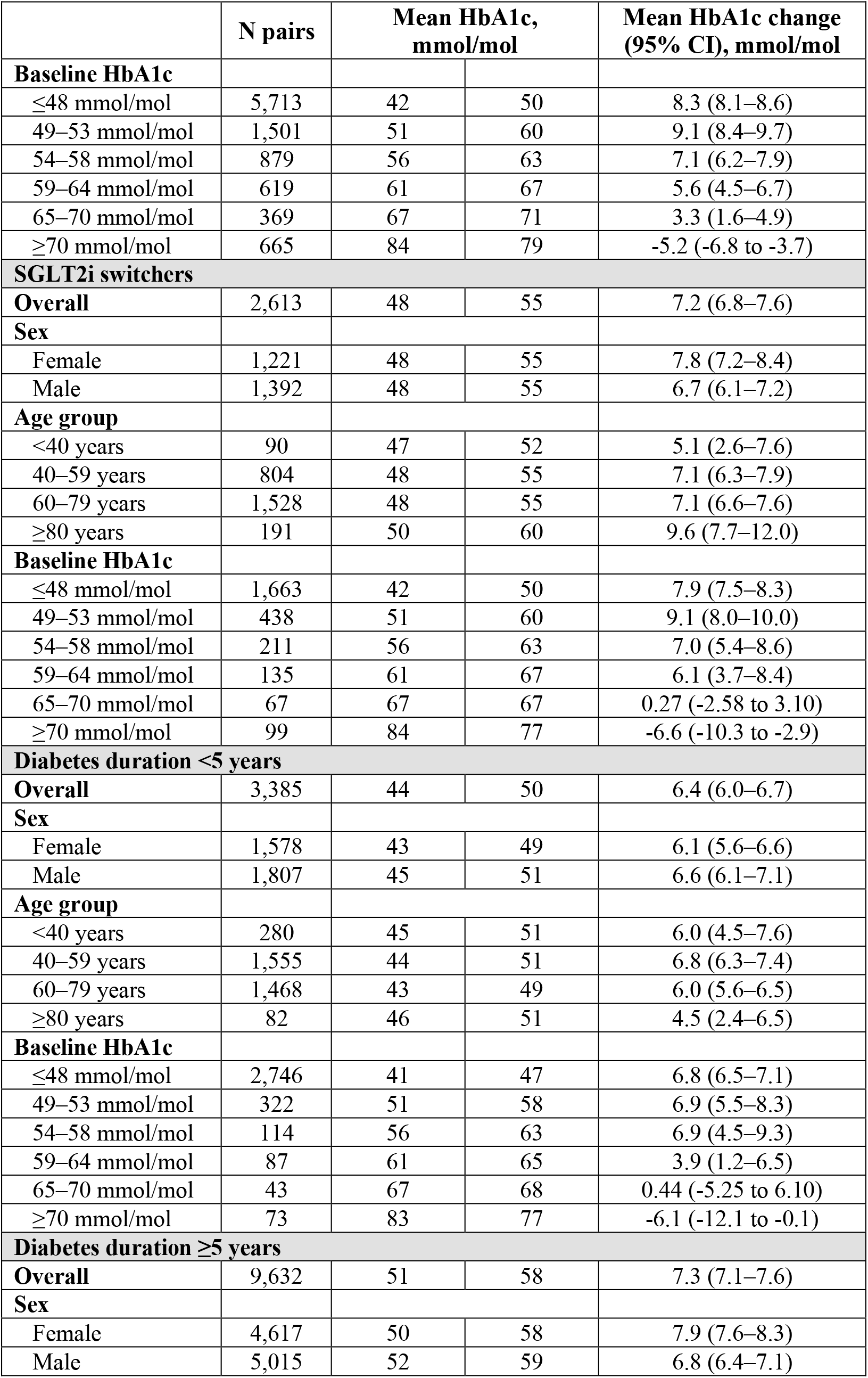
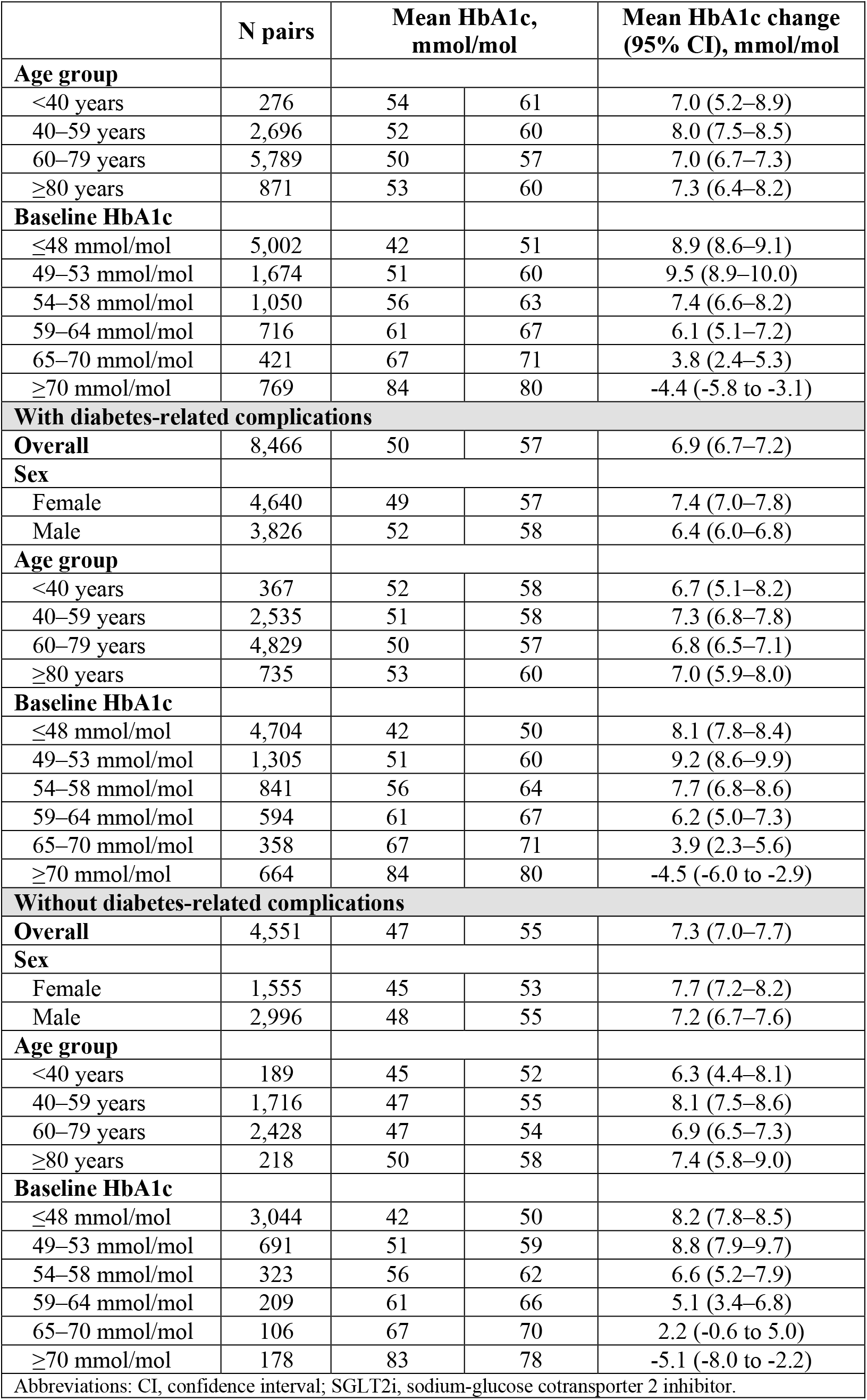
Within-individual change in HbA1c using alternative post-treatment definitions, including the first eligible post-HbA1c measurement, the latest post-HbA1c measurement, stockpiling-adjusted estimates, switching to sodium-glucose cotransporter 2 inhibitors, diabetes duration, and complication status.

**Table S7.** Interrupted time series estimates of HbA1c level and trend changes among discontinuers, continuers, and SGLT2i switchers.

|  | **HbA1c, mmol/mol**<br>**(95% CI)** | **p-value** |
| --- | --- | --- |
| **Discontinuers of semaglutide (N = 21,264)** |  |  |
| **Intercept (baseline level)** | 51.8 (51.4 to 52.2) | <0.001 |
| **Pre-intervention trend** | 0.10 (0.04 to 0.16) | 0.003 |
| **Level-change (immediate effect at six months)** | 5.9 (5.1 to 6.7) | <0.001 |
| **Post-intervention trend change** | -0.46 (-0.64 to -0.29) | <0.001 |
| **Effect at 12 months** | 3.1 (2.2 to 4.1) | <0.001 |
| **Continuers of semaglutide (N = 56,488)** |  |  |
| **Intercept (baseline level)** | 53.8 (53.2 to 54.5) | <0.001 |
| **Pre-intervention trend** | -0.14 (-0.23 to -0.05) | 0.004 |
| **Level-change (immediate effect at six months)** | 1.4 (0.2 to 2.6) | 0.023 |
| **Post-intervention trend change** | -0.04 (-0.31 to 0.23) | 0.755 |
| **Effect at 12 months** | 1.2 (-0.2 to 2.6) | 0.092 |
| **Three months excluded (N = 21,264)** |  |  |
| Estimates are not reported* |  |  |
| **SGLT2i switchers (3,704)** |  |  |
| **Intercept (baseline level)** | 48.9 (48.2 to 49.5) | <0.001 |
| **Pre-intervention trend** | 0.02 (-0.06 to 0.11) | 0.578 |
| **Level-change (immediate effect at six months)** | 8.7 (7.6 to 9.8) | <0.001 |
| **Post-intervention trend change** | -0.82 (-1.07 to -0.56) | <0.001 |
| **Effect at 12 months** | 3.8 (2.5 to 5.1) | <0.001 |
| Abbreviations: CI, confidence interval.<br>*Interrupted time series estimates are not reported because the segmented linear regression model does not fit the observed monthly mean HbA1c data. The observed data are shown in Figure S4. |  |  |

**Figure S1.**
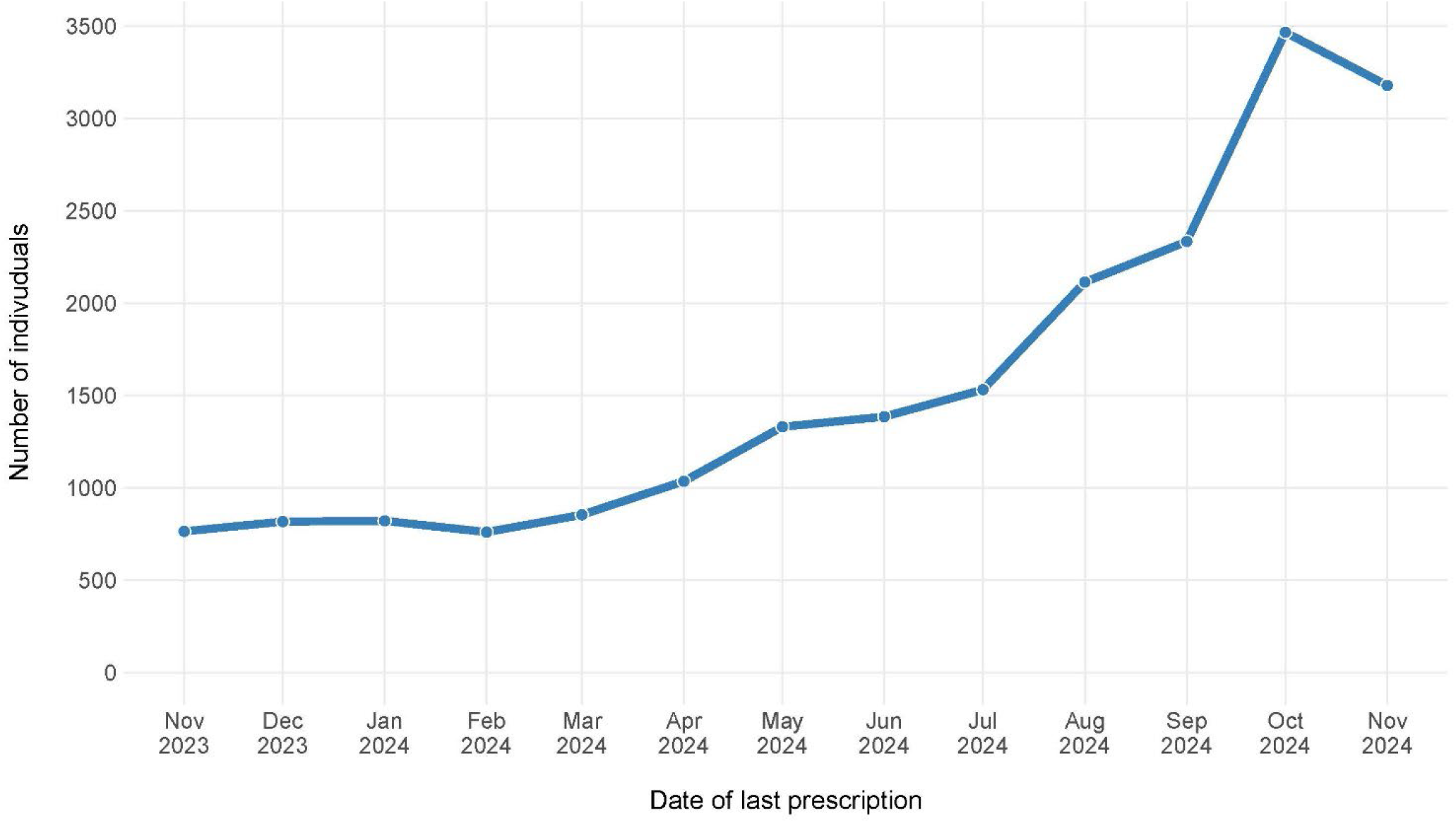
Exploratory analysis of monthly discontinuations among semaglutide users from November 2023 to November 2024, defined as the last fill of semaglutide during this period, with no subsequent GLP-1RA fills in the six months following 25 November 2024.

**Figure S2.**
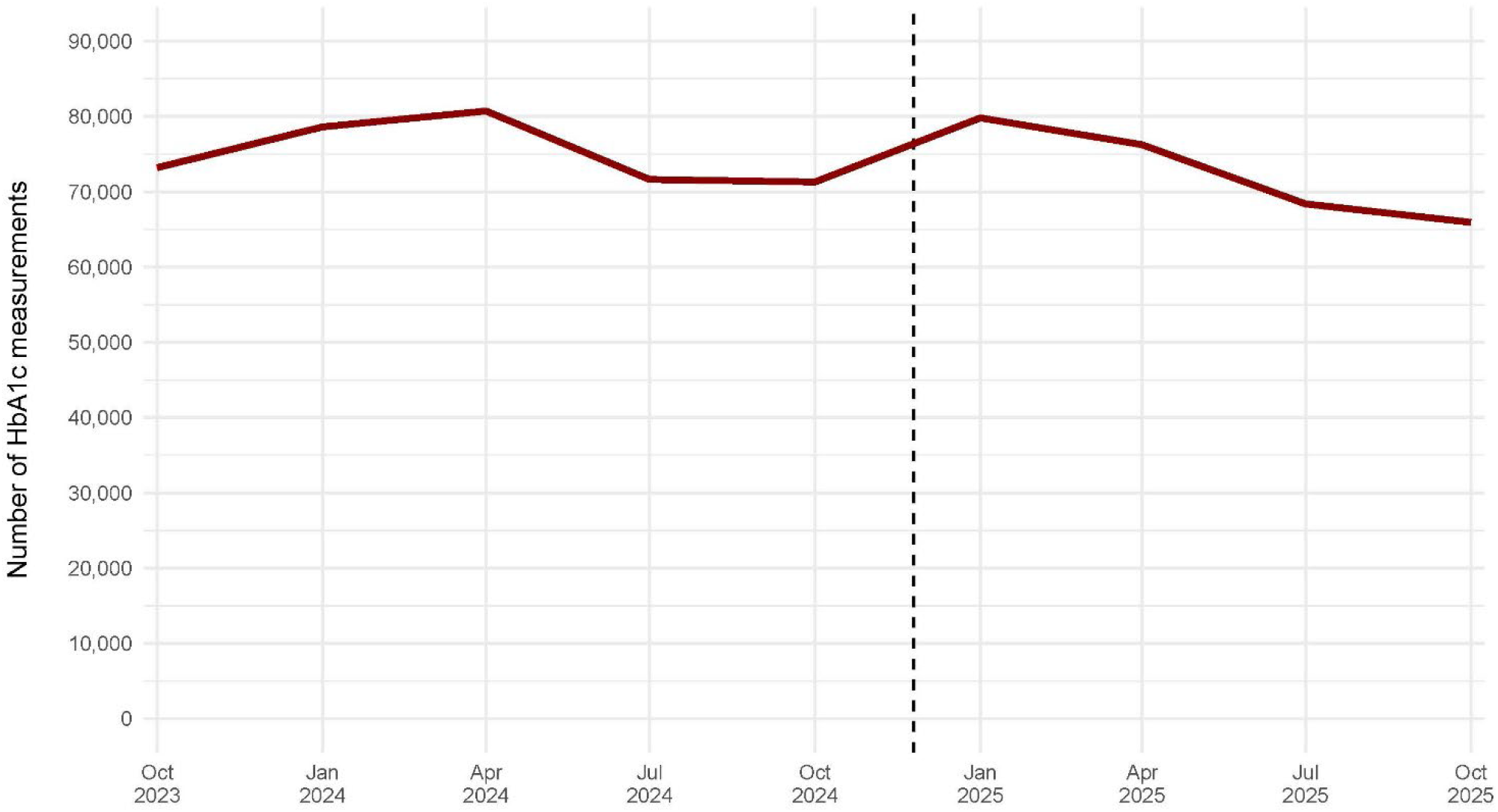
Quarterly number of HbA1c measurements in adults using injectable semaglutide for diabetes management from October 2023 to October 2025, with the timing of the policy change indicated with the dashed line.

**Figure S3.**
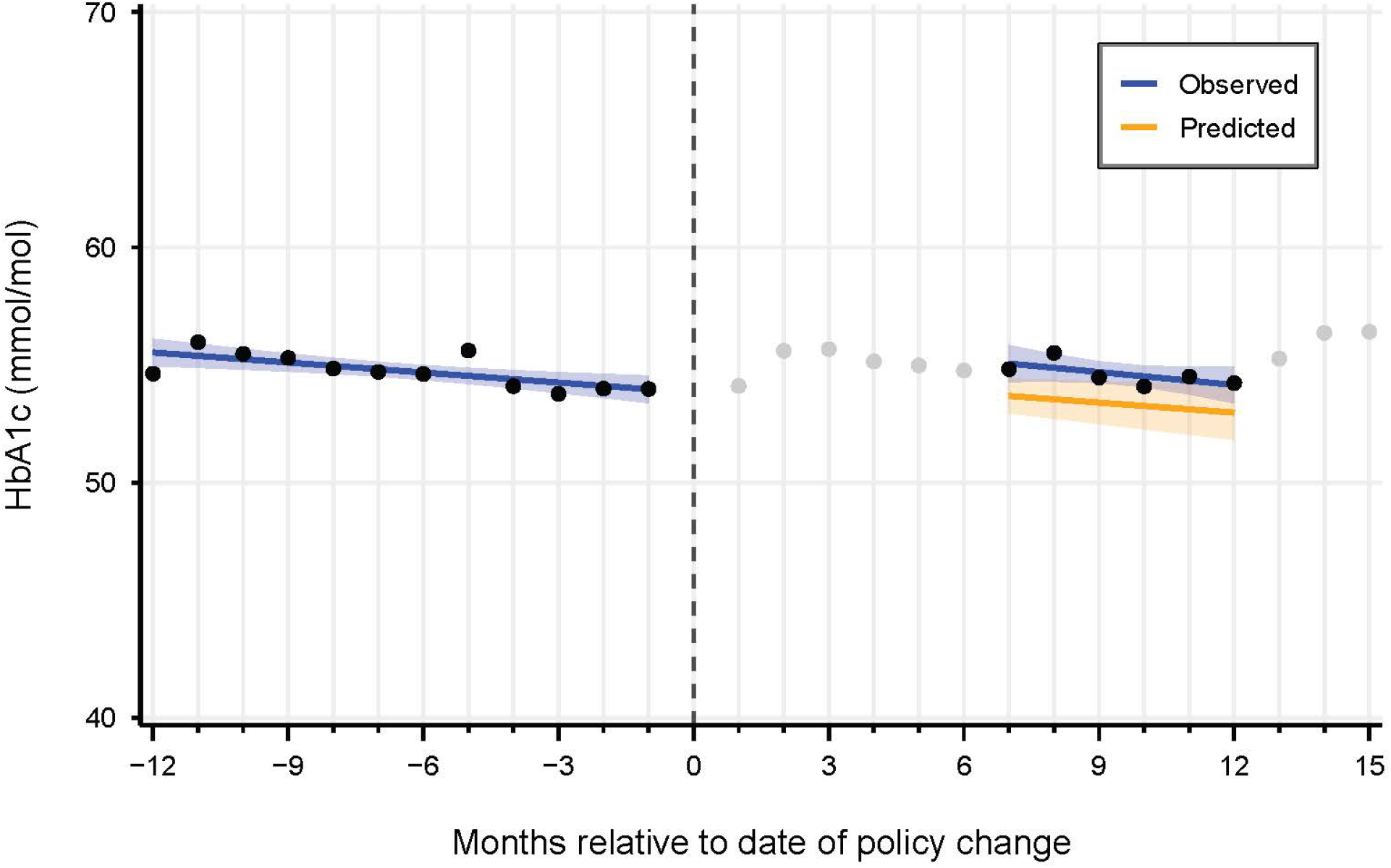
Interrupted time series of monthly mean HbA1c 12 months before and 6–12 months after the policy change (i.e., 25 November 2024) among continued semaglutide users (N = 56,488). The black points represent observed HbA1c values included in the analysis. The blue line shows the fitted linear model based on the observed data points. The orange line is the counterfactual prediction. The grey points represent observed HbA1c values not included in the analysis. Shaded areas indicate 95% confidence intervals.

**Figure S4.**
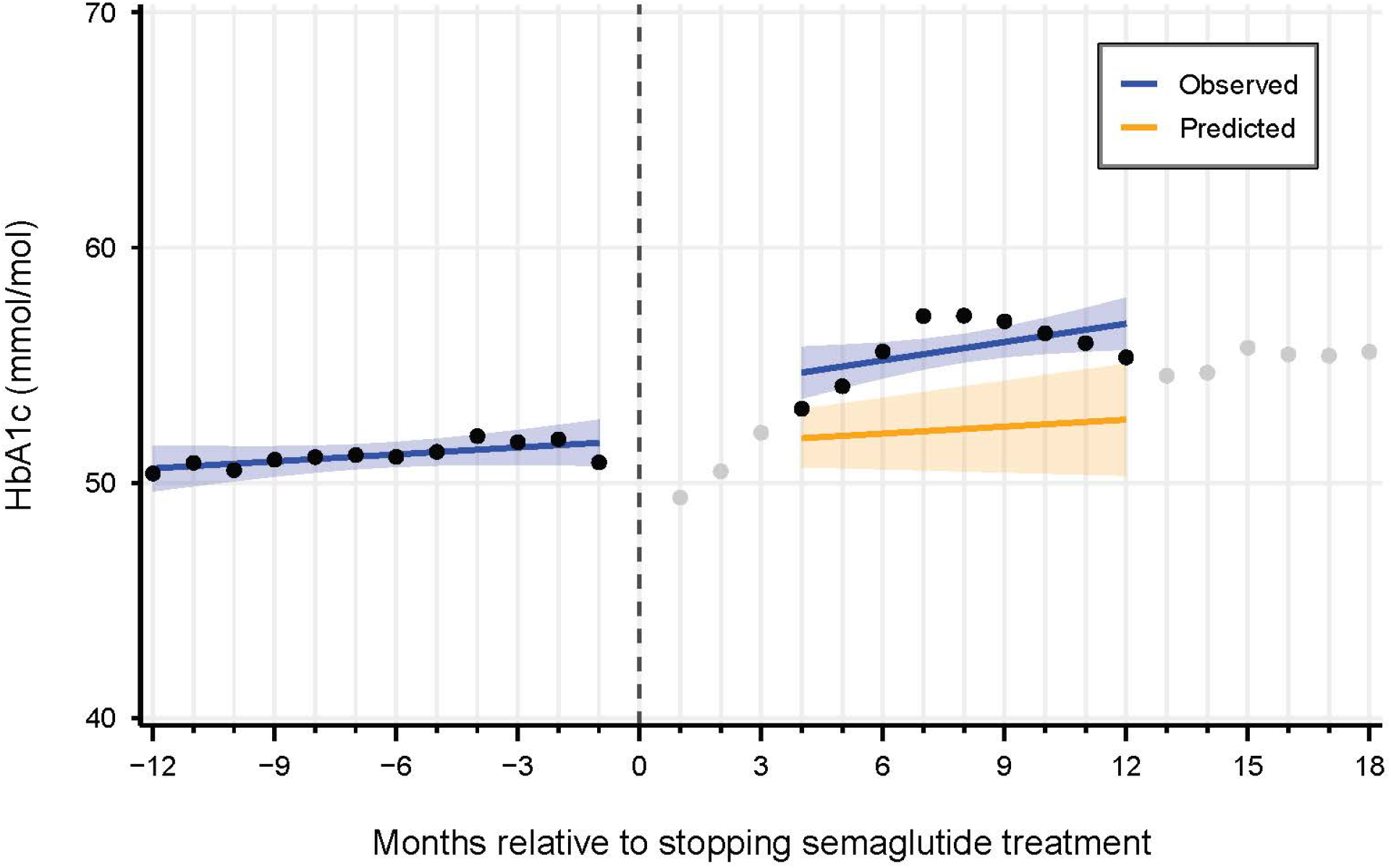
Interrupted time series of monthly mean HbA1c 12 months before and 3–12 months after semaglutide discontinuation (N = 21,264). The black points represent observed HbA1c values included in the analysis. The blue line shows the fitted linear model based on the observed data points. Because of poor model fit, segmented regression estimates are not reported. The orange line is the counterfactual prediction. The grey points represent observed HbA1c values not included in the analysis. Shaded areas indicate 95% confidence intervals.

**Figure S5.**
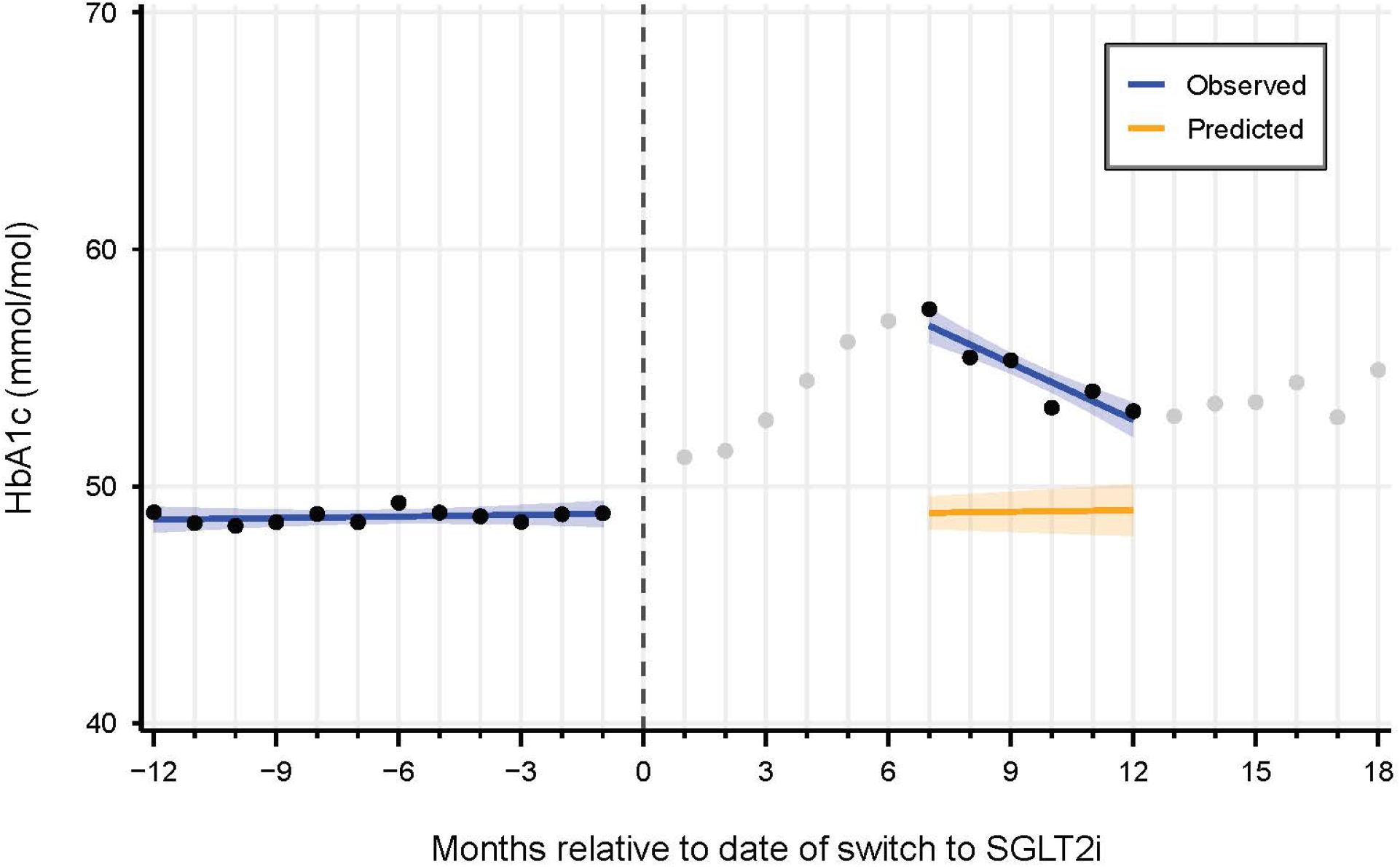
Interrupted time series of monthly mean HbA1c 12 months before and 6–12 months after switch to a sodium-glucose cotransporter 2 inhibitor (SGLT2i) (N = 3,704). The black points represent observed HbA1c values included in the analysis. The blue line shows the fitted linear model based on the observed data points. The orange line is the counterfactual prediction. The grey points represent observed HbA1c values not included in the analysis. Shaded areas indicate 95% confidence intervals.

